# Exploring the longitudinal associations of functional network connectivity and psychiatric symptom changes in youth

**DOI:** 10.1101/2022.11.26.22282787

**Authors:** Lorenza Dall’Aglio, Fernando Estévez-López, Mónica López-Vicente, Bing Xu, Oktay Agcaoglu, Elias Boroda, Kelvin O. Lim, Vince D. Calhoun, Henning Tiemeier, Ryan L. Muetzel

**Author notes:** **CORRESPONDANCE** Henning Tiemeier, M.D., Ph.D., Address: 677 Huntington Avenue, Boston, MA, 02115, USA.

## Abstract

**Background:** Functional connectivity has been associated with psychiatric problems, both in children and adults, but inconsistencies are present across studies. Prior research has mostly focused on small clinical samples with cross-sectional designs.

**Methods:** We adopted a longitudinal design with repeated assessments to investigate associations between functional network connectivity (FNC) and psychiatric problems in youth (9- to 17-year-olds) from the general population. The largest single-site study of pediatric neurodevelopment was used: Generation R (*N* = 3,131). Psychiatric symptoms were measured with the Child Behavioral Checklist as broadband internalizing and externalizing problems, and its eight specific syndrome scales (e.g., anxious-depressed). FNC was assessed with two complementary approaches. First, static FNC (sFNC) was measured with graph theory-based metrics. Second, dynamic FNC (dFNC), where connectivity is allowed to vary over time, was summarized into 5 states that participants spent time in. Cross-lagged panel models were used to investigate the longitudinal bidirectional relationships of sFNC with internalizing and externalizing problems. Similar cross-lagged panel models were run for dFNC.

**Results:** Small longitudinal relationships between dFNC and certain syndrome scales were observed, especially for baseline syndrome scales (i.e., rule-breaking, somatic complaints, thought problems, and attention problems) predicting connectivity changes. However, no association between any of the psychiatric problems (broadband and syndrome scales) with either measure of FNC survived correction for multiple testing.

**Conclusion:** We found no or very modest evidence for longitudinal associations between psychiatric problems with dynamic and static FNC in this population-based sample. Differences in findings may stem from the population drawn, study design, developmental timing and sample sizes.

---

Psychiatric problems tend to arise in early life (Solmi et al., 2022), with childhood and adolescence being considered key developmental windows in which substantial changes in behavioral issues emerge (Galván, 2017). Neurobiological differences are among the proposed mechanisms that determine psychiatric problem occurrence. The neurodevelopmental changes taking place in youth have made neurobiology a prime candidate for investigations into psychiatric disorders’ etiology (Vijayakumar et al., 2018). Specifically, a large body of literature has shown, albeit inconsistently, relationships between psychiatric problems and brain functional connectivity (FC) and its network analog, functional network connectivity (FNC), in childhood and adolescence (Canario et al., 2021).

FNC refers to the temporal correlation among functional communities in the brain, also called resting-state networks (van den Heuvel & Sporns, 2013). Brain FNC can be assessed with resting-state functional magnetic resonance imaging (rs-fMRI). This sequence can be used to quantify correlated spontaneous low-frequency fluctuations in the BOLD signal across brain networks (Biswal et al., 1995). FNC can be measured both in a static and dynamic manner. Static FNC (sFNC) allows the capturing of connectivity patterns across the brain, such as average connectivity in a given network. Dynamic FNC (dFNC) extends this by examining the dynamicity of brain networks, meaning that it allows for correlational patterns across regions to vary over time during imaging assessment (Allen et al., 2014). Importantly, FNC has been shown to reliably represent brain development (Allen et al., 2011; López-Vicente et al., 2021). However, studies leveraging either rs-fMRI approach to better understand psychopathology face numerous challenges, including poor replicability of clinical findings (Griffanti et al., 2016).

Prior literature investigated FNC patterns, at both static and dynamic levels, in several psychiatric disorders. Differences in FNC between cases and controls have been identified, although in different areas or for different properties, meaning that findings remain inconsistent or difficult to compare (Oldehinkel et al., 2013). For sFNC, several clinical studies found disrupted network properties for major depressive disorder (Wise et al., 2017), autism spectrum disorder (Sha et al., 2022), and attention-deficit/hyperactivity disorder (Saad et al., 2017). Results suggest differences in the efficiency of the sharing of information across networks, at the global or local levels (global and local efficiency). For dFNC, differential patterns were present for major depression (Wise et al., 2017; Wu et al., 2019; Zhi et al., 2018), schizophrenia (Damaraju et al., 2014; Ma et al., 2014; Sendi et al., 2021), and neurodevelopmental disorders (de Lacy & Calhoun, 2018). Patients suffering from mental illness generally spent more time in states characterized by inefficient connectivity or global dysconnectivity. Moreover, some studies found that psychiatric patients spent more time in states related to self-focused thinking, less in states for positive emotions, and had higher fluctuations across states (Ma et al., 2014; Wise et al., 2017; Wu et al., 2019).

While this body of literature suggests that brain FNC may further our understanding of psychiatric problems, interpretation is often hampered by *cross-sectional study designs*. Longitudinal approaches are key to shedding further light on the relationships between FNC and behavioral problems and can inform on the temporal directionality of associations, if repeated measurements are available. Further, prior literature has accounted for a few selected *confounders* (e.g., age, sex), but additional factors may be at play in associations between brain functioning and psychiatric issues (e.g., socioeconomic status) (Dall’Aglio, Kim, et al., 2022). Additionally, previous studies mostly used *diagnostic data* in *clinical samples*. However, psychiatric problems likely exist on a continuum (Garvey et al., 2016), highlighting the need for exploring the full extent of psychiatric symptoms and using population-based samples. Such explorations may be particularly beneficial in key *developmental periods* like adolescence, during which several psychiatric problems onset or exacerbate and substantial neurodevelopmental changes occur (Solmi et al., 2022; Vijayakumar et al., 2018). Lastly, examinations of FNC in relation to psychiatric problems in youth have been mostly limited to *static* approaches (Canario et al., 2021; López-Vicente et al., 2021). Adopting dynamic approaches to study FNC may provide novel information regarding the connectivity pattern changes occurring with psychiatric problems.

Here, we explored the longitudinal relationships between brain connectivity and psychiatric symptoms, as measured by static and dynamic FNC approaches, in a large (*N* = 3,131), single-site, population-based sample of adolescents, leveraging repeated rs-fMRI and behavioral assessments.

## METHODS

### Participants

This longitudinal study was embedded within the Generation R Study, a prospective birth cohort from fetal life up until adolescence (Kooijman et al., 2016), which is conducted in Rotterdam, the Netherlands (one study site). Ethical approval was obtained from the Medical Ethics Committee of Erasmus MC, University Medical Center Rotterdam. All parents provided written informed consent and children provided assent (younger than 12 years) or consent (12 years or older).

Of the Generation R Study cohort (*N* = 9,901), we used data from children with assessments at ages 10- or 14-years (age range: 9- to 17-year-olds). Children with data on psychiatric problems (internalizing or externalizing) and functional connectivity at one assessment were included. In total, 3,767 children were imaged at either assessment, with 1,037 presenting data for both assessments. After applying the exclusion criteria, consisting of clinically relevant incidental findings, and poor image quality (see *Image quality control*), we obtained a sample of 3,296 children. For each sibling or twin in the sample, one was randomly included to prevent clustering. We obtained a final sample of 3,131 children from the general population with connectivity and psychiatric data.

### Measures

#### Image acquisition

Images were obtained on a study-dedicated 3T scan (GE Discovery MR750w MRI System, General Electric, Milwaukee, WI, United States) with an eight-channel head coil. Rs-fMRI data were collected with an interleaved gradient recalled axial-echo planar imaging sequence (*T*_R_ = 1,760 ms, *T*E = 30 ms, flip angle = 85°, matrix = 64 × 64, FOV = 230 mm × 230 mm, slice thickness = 4 mm) for a total of 5 min. 52 s. per child (White et al., 2018). Participants were instructed to keep their eyes closed and stay awake.

#### Image pre-processing

Pre-processing of the images was performed with the FMRIPrep pipeline (v. 20.1.1 singularity container) (Esteban et al., 2019). FMRIPrep involves *(i)* volume realignment for motion from rotation and translation, *(ii)* slice-timing correction, and *(iii)* inter-subject registration. Spatial normalization (ICBM 152 non-linear asymmetrical template v. 2009c) (Fonov et al., 2009) was conducted with non-linear registration (antsRegistration tool, ANTs v2.1.0) (Avants et al., 2008). Functional data were resampled to 3mm x 3mm x 3mm isotropic voxels. Volume realignment provided us with the time series for calculating the first temporal derivatives of the six base motion parameters (three rotations, three translations) as well as quadratic terms (six for the base parameters and six for the temporal derivatives). We obtained a total of 24 head motion parameters, which were used as confound regressors before connectivity analysis (Satterthwaite et al., 2013).

#### Image quality control

We first excluded scans with high motion, based on a mean framewise displacement higher than 0.25 mm or more than 20% of the volumes with a framewise displacement higher than 0.2 mm (Parkes et al., 2018). Second, visual inspections were performed for image co-registration, major artifacts, and whole-brain coverage. Problematic images, based on these criteria, were excluded (López-Vicente et al., 2021). In a sensitivity analysis, more stringent quality control was applied, where a mean framewise displacement ≤0.11 mm cutoff was used for exclusion (see *Sensitivity analyses*).

#### Static functional network connectivity

We summarized static FNC characteristics using a graph theory-based approach representing the topological architecture of brain networks. Graph theory provides information on functional networks across the whole brain and on how networks may relate to each other. This is in line with the evidence that FC is organized into several complex networks used to integrate and segregate information (Fornito et al., 2016). Fifty-one reference components from the Dev-CoG developmental imaging study were used to parcellate the functional scans (Agcaoglu et al., 2019, 2020). The 51x51 correlation matrices were generated by calculating Pearson correlations between the time series and were Fisher r-to-z transformed to obtain normal distributions. The brain connectivity toolbox was then used (python version) (La Plante, 2022; Rubinov & Sporns, 2010). Matrix diagonals were excluded and the lower half of the matrix was extracted. Continuous values were used (i.e., weighted rather than threshold approach). The graph measures calculated included the clustering coefficient, modularity, characteristic path length, and global efficiency. The clustering coefficient indicates the extent to which neighboring nodes connect to each other (i.e., are clustered together) (Berlot et al., 2016). Modularity measures the extent to which networks can be partitioned into segregated modules or communities. Characteristic path length assesses the average number of steps in the shortest paths connecting each pair of nodes. Global efficiency is the inverse of characteristic path length and is considered an indication of the capacity for integrated processing and parallel information transfer (Berlot et al., 2016).

#### Dynamic functional network connectivity

Dynamic FNC techniques allow to capture changes in connectivity across brain areas during the assessment. This is important as differences in functional activity throughout the MRI scanning procedure have been shown (Allen et al., 2014). dFNC is used to identify different functional connectivity states or configurations of a participant, and summary measures, such as the time spent in each state (mean dwell time (MDT)) and the number of transitions across states (NT), can be calculated. Data on dFNC were previously obtained and described for the Generation R Study (López-Vicente et al., 2021). Briefly, the Group ICA of fMRI Toolbox (GIFT) software was used (GroupICAT v4.0b) (Calhoun et al., 2001; Calhoun & Adalı, 2012). dFNC was calculated on the subject-specific time courses using a sliding window approach. A tapered windows of 25 TR (44s) in steps of 1 TR were used. A 3 TR alpha parameter of the Gaussian sliding window was leveraged (Allen et al., 2014; Qin et al., 2015). We obtained 171 FNC windows per person. Covariance was estimated using regularized inverse covariance matrices with a graphical LASSO framework (Friedman et al., 2008; Smith et al., 2011). To ensure sparsity, an L1 norm constraint on the matrices was applied. With cross-validation, we evaluated the log-likelihood of unseen data for each subject/visit to optimize the regularization parameter. K-means clustering of the 171 dFNC windows was used to derive five states (López-Vicente et al., 2021). Three of these states were modularized with components of intra- and inter-network connectivity while two were non- or partially modularized. More specifically, state 1 is a modularized state presenting negative interconnectivity between subcortical and sensorimotor networks (López-Vicente et al., 2021), state 2 was default mode/sensorimotor network modularized while state 3 was default mode network modularized. State 4 and 5 were non-modularized and partially modularized, respectively. States can be visualized in **Figure S1**, with more information being available in prior literature (López-Vicente et al., 2021). The MDT spent in a given state and the NT across states per participant and assessment were then calculated.

#### Psychiatric problems

We measured psychiatric problems with the school-age version of the Child Behavioral Checklist (CBCL) (Achenbach & Rescorla, 2001). The CBCL was completed by the primary caregiver to assess children’s psychiatric symptoms dimensionally at both the age 10 and age 14 assessments. The CBCL is considered a reliable and valid questionnaire, generalizable across societies and nationalities (Achenbach & Rescorla, 2001; Achenbach et al., 2017; Rescorla et al., 2007). Following the ASEBA protocol (Achenbach & Rescorla, 2001), scores were calculated by averaging relevant items and allowing a maximum of 25% of missing items per participant. Raw scores were square-root transformed to address non-normality. We used the broadband measures of psychopathology (internalizing and externalizing symptoms), as well as narrow-band ones (8 syndrome scales: attention, thought, social problems, somatic complaints, anxious-depressed, withdrawn-depressed, aggressive, and rule-breaking behaviors).

### Covariates

Covariates included child demographic characteristics (age, sex, and national origin), highest achieved maternal education, and perceived pubertal status. Parental national origin was assessed through questionnaires and summarized into three categories: Dutch, non-Dutch European, and non-European. Maternal education was measured with self-reported questionnaires and categorized into low (no/primary education), intermediate (secondary school, vocational training), and high (Bachelor’s degree/University). The perceived pubertal stage was assessed at age 14 years based on the Pubertal Development Scale, as an average score of the non-specific and sex-specific items for each sex (as assigned at birth) (Carskadon & Acebo, 1993; Dall’Aglio, Xu, et al., 2022).

### Statistical analyses

#### Main analyses

All statistical analyses were exploratory and performed in the R Statistical Software (version 4.0.1). All scripts will be publicly available online https://github.com/LorenzaDA/rsfMRI_psychiatry_youth_GenR.

To explore the relations of FNC with psychiatric symptoms in youth, we ran the following analyses for static and dynamic FNC measures separately. First, we used cross-lagged panel models to examine the longitudinal relations of connectivity with internalizing and externalizing problems, modeling change from late childhood to early adolescence. Internalizing and externalizing problems were each examined. Overall, 4 measures of static connectivity (the clustering coefficient, global efficiency, modularity, and characteristic path length), and 6 measures of dynamic connectivity (MDT at 5 states and the NTs across states) were examined in association with internalizing/externalizing problems. Multiple testing correction was applied for each set (i.e., dynamic and static FNC separately) using the false discovery rate method (FDR, Benjamini-Hochberg).

Cross-lagged panel models allow for the examination of associations between two repeatedly assessed variables simultaneously. Of the coefficients estimated within cross-lagged panel models, which include lagged effects, covariances, and autoregressive coefficients, here we focused on lagged paths. Lagged effects provide information on whether *(i)* internalizing/externalizing problems at baseline predict changes in FNC over time and *(ii)* FNC at baseline predicts changes in internalizing/externalizing problems over time. The *Lavaan* package was used (Rosseel, 2012). The conceptual model is shown in **Figure S2**. Missing values were imputed within *lavaan* with the full-information maximum likelihood approach, meaning the sample for all analyses was of 3,131 children.

#### Non-response analysis

We conducted a non-response analysis in which we compared non-participating children eligible for the assessments at the age of 10 years to the children in the analyses, based on covariates (parental education, self-perceived puberty, child national origin, sex). Independent samples t-tests and chi-square tests were used for continuous and categorical variables respectively.

#### Sensitivity analyses

We conducted several sensitivity analyses to further explore the results and to evaluate the robustness of our findings. First, all cross-lagged panel models were re-run with the eight CBCL syndrome scales, to explore patterns of connectivity for more specific measures of psychopathology. Multiple testing correction was applied on static and dynamic FNC separately, across all syndrome scales. Second, we evaluated whether the main models fit similarly to males and females, with likelihood ratio tests. More specifically, models grouped by sex where regression coefficients were allowed to differ for boys and girls, were contrasted with models grouped by sex where regression coefficients were restricted to equal across sexes. Third, to ensure that results were not dependent on the motion quality control procedures we used, we rerun our main analyses using a more stringent threshold for framewise displacement (≤0.11 mm instead of 0.25 mm), by excluding participants who had a score higher than the top 20% of framewise displacement values in our cohort (*N* for motion sensitivity analysis = 2,971).

## RESULTS

### Sample characteristics

We included 3,131 individuals with rs-fMRI and CBCL data at either the first (T1) or second assessment (T2). An approximately equal proportion of males and females was included (females *n* = 1,634 (52%)). Children were, on average, 10 years old at T1 and 14 years old at T2 (age ranges: 9-13 years (T1), 13-17 years (T2)) (**Table S1**). Non-response analyses indicated that participants represented the full cohort at T1 in terms of sex, maternal education, national origin, and self-perceived pubertal status (**Table S2**).

### Static connectivity

Static FNC properties are shown in **Figure S3** and **Table S1**. We tested the longitudinal relationships between internalizing problems and sFNC (characteristic path length, global efficiency, clustering coefficient, modularity) by modeling change over time with cross-lagged panel models (**Figure S2**). The same models were run for externalizing problems. The models fit the data well for both broadband scales, as evidenced by the absolute and relative model fit indices in **Table S3**. We did not identify a statistically significant relationship between internalizing problems and network properties in either temporal direction, i.e., sFNC was not associated with changes in psychiatric symptoms (**Table 1**) nor were psychiatric symptoms with changes in sFNC (**Table 2**). Similar results were observed for externalizing problems (**Tables 1-2**). Estimates for longitudinal effects ranged from -0.036 to 0.028, with standard errors ranging from 0.018 to 0.019 (**Tables 1-2**). Analogous results were found when using more stringent motion corrections (**Table S4**). Moreover, we found no evidence of sex differences in the relationships between static connectivity and internalizing or externalizing problems, as shown by the lack of significant improvement in model fit when regression coefficients were allowed to vary vs. be equal across sexes (**Table S5**).

**Table 1.**
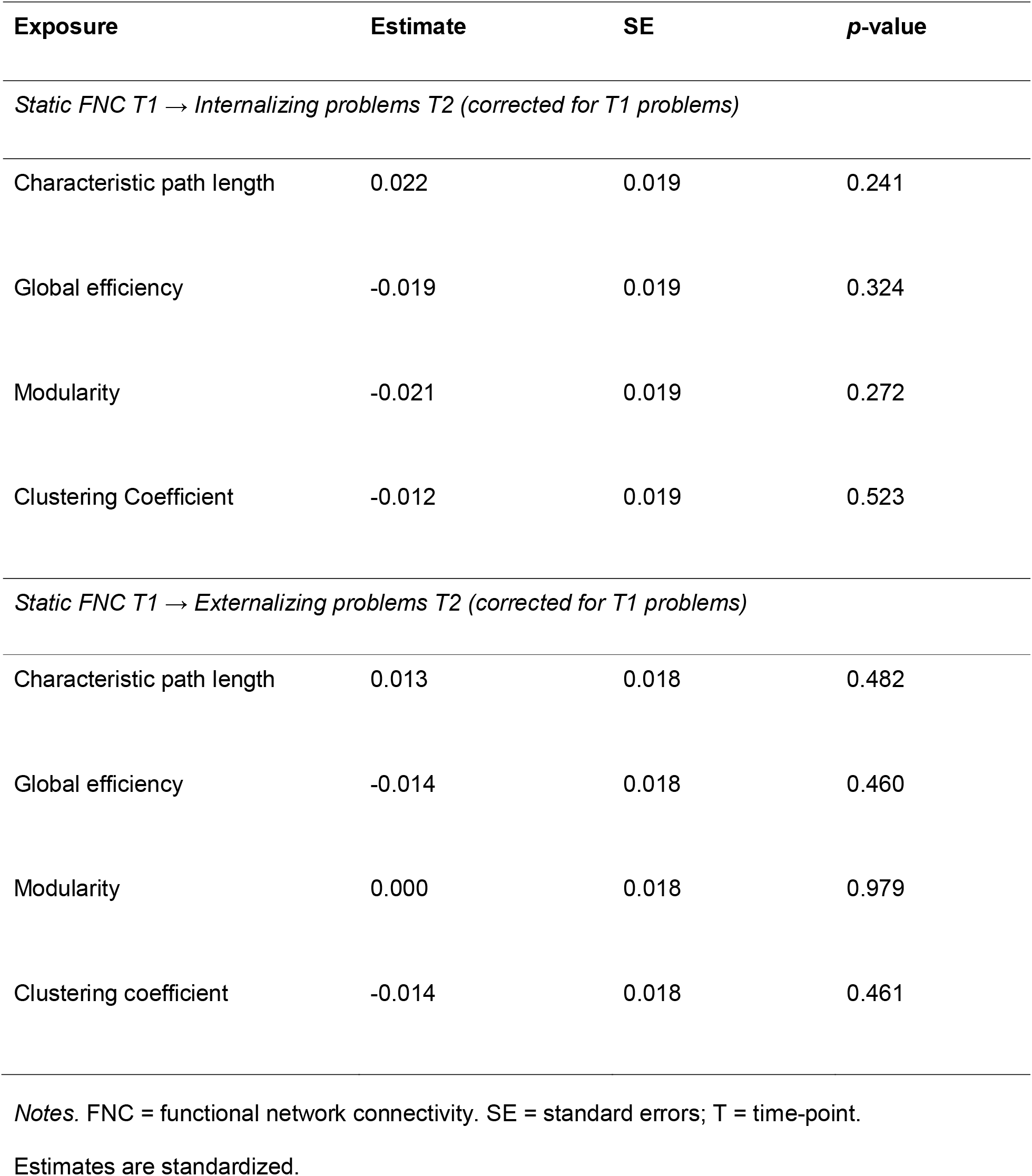
Longitudinal relationships of baseline static functional network connectivity with internalizing and externalizing problems changes from late childhood to early adolescence

| **Exposure** | **Estimate** | **SE** | **p-value** |
| --- | --- | --- | --- |
| *Static FNC T1 → Internalizing problems T2 (corrected for T1 problems)* |  |  |  |
| Characteristic path length | 0.022 | 0.019 | 0.241 |
| Global efficiency | -0.019 | 0.019 | 0.324 |
| Modularity | -0.021 | 0.019 | 0.272 |
| Clustering Coefficient | -0.012 | 0.019 | 0.523 |
| *Static FNC T1 → Externalizing problems T2 (corrected for T1 problems)* |  |  |  |
| Characteristic path length | 0.013 | 0.018 | 0.482 |
| Global efficiency | -0.014 | 0.018 | 0.460 |
| Modularity | 0.000 | 0.018 | 0.979 |
| Clustering coefficient | -0.014 | 0.018 | 0.461 |

*Notes.* FNC = functional network connectivity. SE = standard errors; T = time-point.
Estimates are standardized.

**Table 2.** Longitudinal associations of baseline internalizing and externalizing problems with static functional network connectivity changes from late childhood to early adolescence.

| **Model** | **Estimate** | **SE** | **p-value** |
| --- | --- | --- | --- |
| *Internalizing problems T1 → static FNC T2 (corrected for T1 FNC)* |  |  |  |
| Characteristic path length | 0.028 | 0.024 | 0.245 |
| Global efficiency | -0.027 | 0.024 | 0.276 |
| Modularity | -0.017 | 0.024 | 0.474 |
| Clustering Coefficient | -0.020 | 0.025 | 0.423 |
| *Externalizing problems T1 → static FNC T2 (corrected for T1 FNC)* |  |  |  |
| Characteristic path length | -0.009 | 0.024 | 0.706 |
| Global efficiency | 0.006 | 0.025 | 0.797 |
| Modularity | -0.036 | 0.024 | 0.138 |
| Clustering Coefficient | 0.011 | 0.025 | 0.647 |

*Note.* FNC = functional network connectivity; SE = standard error; T = time point. Estimates are standardized.

Longitudinal relations of static connectivity were also tested in relation to *specific* psychiatric problems (i.e., 8 syndrome scales), to explore whether associations were present at a more fine-grained level. We did not identify any longitudinal associations, in either temporal direction, between specific psychiatric problems and static connectivity (**Table S6**). Estimates ranged from -0.047 to 0.038 and standard errors from 0.018 to 0.025 (**Table S6**).

### Dynamic FNC

Dynamic FNC properties are shown in **Figure 1, Panel A** and **Table S1**. We tested the longitudinal bidirectional relationship of internalizing problems with dFNC for MDT in 5 connectivity states and the number of transitions across states by modeling change with cross-lagged panel models (**Figure S2**). The same models were run for externalizing problems. The models of dFNC with each broadband scale fit the data well, as shown in **Table S7**. In the analyses of baseline dFNC with changes in internalizing problems (brain → behavior), no significant association was identified, before or after multiple testing corrections (*p*_FDR_ < 0.05) (**Table 3**). Largely similarly negative results were found for dFNC as determinant of changes in externalizing problems (**Table 3**), although MDT in state 5 at baseline nominally predicted changes in externalizing problems (estimate = -0.048; SE = 0.018; *p*-value = 0.009). This association did not surpass multiple testing corrections (*p*_FDR_ = 0.212) (**Table 3**), but it was robust to stricter motion control in a sensitivity analysis (estimate = -0.048; SE = 0.019; *p*-value = 0.010) (**Table S8**).

**Figure 1.**
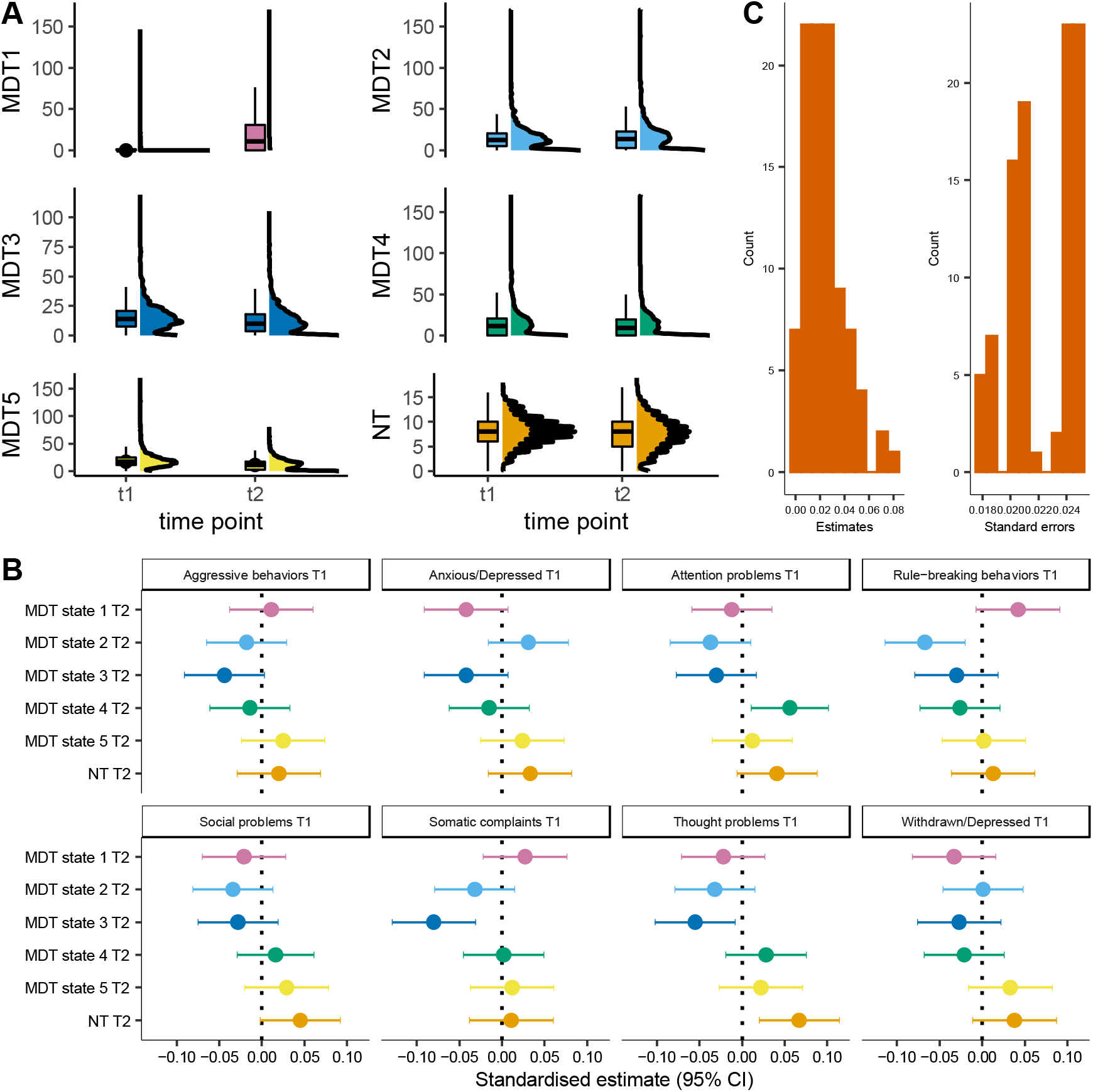
Baseline dynamic functional network connectivity and change in specific psychiatric symptoms from late childhood to early adolescence. *Note*. CI = confidence interval; MDT = mean dwell time for a given state; NT = number of transitions; T = time-point. All estimates are standardized. **A**. Raincloud plots for dynamic functional network connectivity measures across time-point (t1 = age 10 assessment; t2 = age 14 assessment). **B**. Relationships between specific psychiatric symptoms (i.e., 8 syndrome scales) at baseline, in late childhood (T1), with change in dFNC from late childhood to early adolescence (T2). **C**. Histograms of the distribution of standardized absolute estimates and standard errors for all associations of syndrome scales with dFNC.

**Table 3.** Longitudinal associations of baseline dynamic functional network connectivity with internalizing and externalizing problem changes from late childhood to early adolescence.

| **Model** | **Estimate** | **SE** | **p-value** |
| --- | --- | --- | --- |
| *Dynamic FNC T1 → Internalizing problems T2 (corrected for T1 problems)* |  |  |  |
| Mean dwell time state 1 | 0.014 | 0.019 | 0.457 |
| Mean dwell time state 2 | -0.013 | 0.019 | 0.493 |
| Mean dwell time state 3 | 0.007 | 0.019 | 0.719 |
| Mean dwell time state 4 | 0.029 | 0.019 | 0.128 |
| Mean dwell time state 5 | -0.014 | 0.019 | 0.465 |
| Number of transitions | 0.006 | 0.019 | 0.758 |
| *Dynamic FNC T1 → Externalizing problems T2 (corrected for T1 problems)* |  |  |  |
| Mean dwell time state 1 | 0.028 | 0.019 | 0.131 |
| Mean dwell time state 2 | 0.014 | 0.018 | 0.448 |
| Mean dwell time state 3 | 0.029 | 0.018 | 0.113 |
| Mean dwell time state 4 | 0.024 | 0.019 | 0.197 |
| Mean dwell time state 5 | -0.048 | 0.018 | 0.009 |

| **Model** | **Estimate** | **SE** | ***p*-value** |
| --- | --- | --- | --- |
| Number of transitions | 0.017 | 0.018 | 0.365 |

*Note.* FNC = functional network connectivity; SE = standard error; T = time-point. Estimates are standardized.

For baseline internalizing problems with changes in dFNC (behavior → brain), a relationship with MDT in state 3 was initially observed (estimate = -0.052; SE = 0.024; *p*-value = 0.034), but this did not survive multiple testing correction. It was also not robust to stricter motion quality control (estimate = -0.040; SE = 0.026; *p*-value = 0.121). We did not detect any longitudinal relationship between baseline externalizing problems with changes in dFNC. Generally, effect sizes were small and ranged from -0.052 to 0.034, with standard errors ranging from 0.019 to 0.025 (**Table 3-4**). Similar results were found when stricter motion control was applied in sensitivity analyses (**Table S8**). No sex differences for internalizing and externalizing problems with MDT and NT in dFNC were identified (**Table S9**).

**Table 4.**
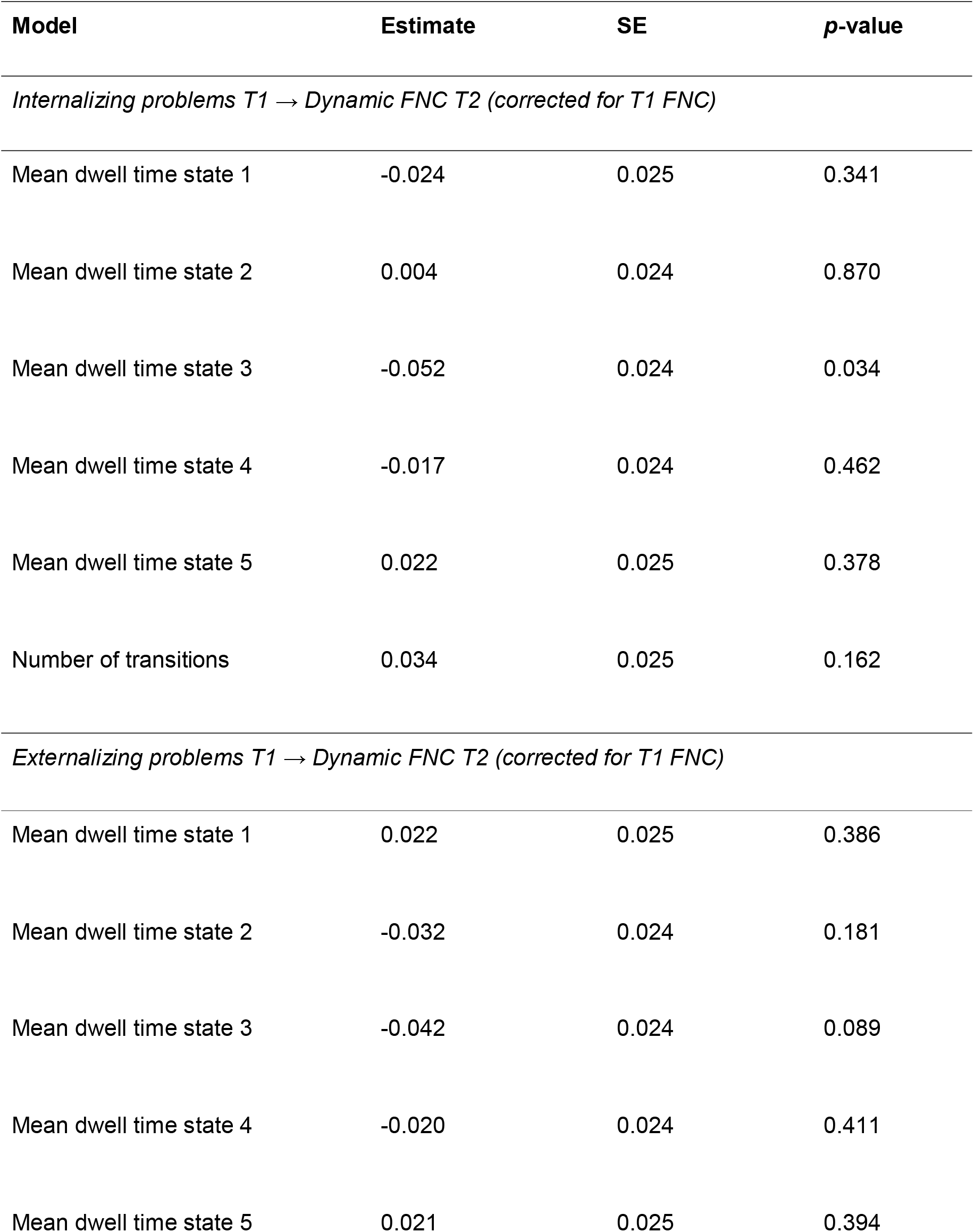
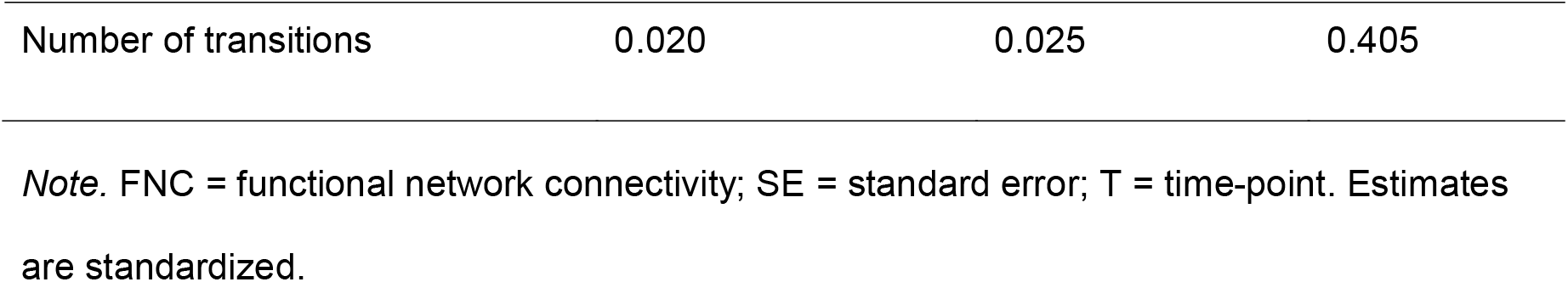
Longitudinal associations of baseline internalizing and externalizing problems with dynamic functional network connectivity changes from late childhood to early adolescence.

Next, the longitudinal relations of dFNC with the specific psychiatric problems (i.e., 8 syndrome scales) were tested. For baseline dFNC with changes in specific psychiatric problems (brain → behavior), we observed that MDT in state 5 determined changes in attention and aggression problems (attention: estimate = -0.056; SE = 0.019; *p*-value = 0.003; *p*_FDR_ = 0.089; aggression: estimate = -0.056; SE = 0.018; *p*-value = 0.002; *p*_FDR_ = 0.089) (**Table S10**). In the analyses of baseline psychiatric symptoms with changes in dFNC (behavior → brain), as shown in **Figure 1, panel B**, we detected several nominal associations. Rule-breaking behaviors at baseline predicted MDT in state 2 over time (estimate = -0.067; SE = 0.024; *p*-value = 0.006; *p*_FDR_ = 0.119), somatic and thought problems for MDT in state 3 (somatic: estimate = -0.080; SE = 0.025; *p*-value = 0.001; *p*_FDR_ = 0.089; thought: estimate = -0.055; SE = 0.024; *p*-value = 0.023; *p*_FDR_ = 0.322), attention problems for MDT in state 4 (estimate = 0.056; SE = 0.023; *p*-value = 0.017; *p*_FDR_ = 0.271), and thought problems predicted change in NT (estimate = 0.067; SE = 0.024; *p*-value = 0.006; *p*_FDR_ = 0.119) (**Table S10**). None of these associations survived multiple testing correction. Overall, median absolute effect sizes were 0.009, while median standard errors were larger, 0.023 (**Figure 1, panel C**).

## DISCUSSION

### Key findings in light of prior literature

This is the largest single-site longitudinal study of static and dynamic functional network connectivity and psychiatric symptoms in youth. Overall, we did not observe any robust longitudinal association between static (general network properties) or dynamic (5 states) FNC with psychiatric problems from age 9 to 17 in the Dutch general population. Some suggestive evidence for longitudinal associations between specific psychiatric problems (rule-breaking, somatic complaints, thought, and attention problems) and dynamic FNC was observed, especially when psychiatric symptoms at baseline predicted changes in dynamic FNC. Yet, none of these associations survived the high multiple testing burden. Relationships must be confirmed in future investigations of specific psychiatric symptoms and changes in FNC dynamicity and will thus be discussed only briefly.

The effect sizes observed here were generally small, based on benchmarks from the Adolescent Brain Cognitive Development (ABCD) Study (Owens et al., 2021). For instance, the magnitude of effects was similar to the cross-sectional relationships of age and prosocial behavior (estimate = 0.01) (Owens et al., 2021). Moreover, for dFNC, our effect sizes were approximately an order of magnitude smaller than the effects of sex or age on dFNC, as previously found in the Generation R Study within the same timeframe (López-Vicente et al., 2021). Further, given the relatively large standard errors we observed, we cannot rule out any true effects may be even smaller. This supports that no or only minor associations are likely present between the brain graph and state-based FNC with psychiatric symptoms during the transition from late childhood to adolescence in this population-based cohort. This is in line with other large studies investigating cross-sectional relationships between psychiatric problems and FNC in ABCD, which is also population-based and sampled 10 to 12-year-olds (Cai et al., 2021; Karcher et al., 2019).

Only a few associations stood out due to their larger effect estimates (**Figure 1, panel B**). These are discussed below. Given the exploratory nature of this study and our high multiple testing burden, they may simply be chance findings and should thus be considered with the utmost care. However, it has been recently suggested that multiple testing corrections may be overly stringent and determine an excess of false-negative results (Marek et al., 2022). A careful discussion of some of these associations may, therefore, be beneficial for future research, with replication being pivotal to evaluating their relevance (Marek et al., 2022).

For static FNC, no relationships were observed before or after multiple testing corrections for any psychiatric problem. For dynamic FNC, we observed nominal relationships for MDT in state 5 with changes in externalizing problems (brain → behavior), which, when further exploring results, seemed to be driven by aggressive symptoms. Associations of MDT in state 5 in late childhood with attention problems in adolescence were also observed. State 5 is a partially modularized state, in which children spend less time as they age, and is marked by sub-modules within networks with distinct connectivity configurations (López-Vicente et al., 2021). Externalizing, aggressive and attention problems generally decrease with age (Verhulst & van der Ende, 1992). In light of this context, our study suggests that higher MDT in state 5 in late childhood might predict smaller decreases in externalizing problems, aggressive symptoms, and attention problems over adolescence.

In the opposite temporal direction (behavior → brain), several specific psychiatric problems predicted changes in dynamic FNC. Rule-breaking behaviors related to MDT in state 2, which is a default-mode/sensorimotor network modularized state relatively stable from late childhood to early adolescence (López-Vicente et al., 2021). Interestingly, cross-sectional associations of externalizing/conduct problems with default-mode network functional connectivity have been previously reported in children and adolescents (Dalwani et al., 2014; Sato et al., 2015). Additionally, somatic complaints and thought problems at baseline predicted smaller changes in MDT in state 3. This is a default-mode network modularized state in which youth spend less time as they grow (López-Vicente et al., 2021). Somatic symptom disorder and schizophrenia were previously associated with differential default mode network connectivity (Broyd et al., 2009; Kim et al., 2019). Higher attention problems associated with larger changes in MDT in state 4, a non-modularized state children spend less time in as they age, especially girls (López-Vicente et al., 2021). Finally, thought problems predicted changes in the number of transitions across states. While typically lower transitions are expected as children age (López-Vicente et al., 2021), with higher thought problems, larger decreases were observed. Importantly, a prior study in the Generation R cohort suggested that behavioral problems influence brain structural connectivity change from early to late childhood (Muetzel et al., 2017), although these results were not found in a study of late childhood into early adolescence (Dall’Aglio, Xu, et al., 2022). As evidence of reverse causality (behavior → brain) is only emerging, it warrants further investigations. Moreover, since dFNC is a novel approach, the literature on the topic remains scarce; future research on its relationship with psychiatric symptoms is particularly desirable.

Importantly, some inconsistencies with prior literature on FNC and psychiatric problems are evident. Altered network topology and dFNC were generally found in individuals with psychiatric disorders, compared to controls (Damaraju et al., 2014; de Lacy & Calhoun, 2018; Ma et al., 2014; Saad et al., 2017; Sendi et al., 2021; Wu et al., 2019; Zhi et al., 2018). Divergent results may stem from differences in several study aspects, such as the population drawn (clinical vs. population-based; age range), study design (cross-sectional vs. longitudinal), and the sample sizes. Previous studies generally focused on *clinical* populations with high levels of psychiatric problems. These often use highly selected groups for comparison, which, compared to the general population, may lead to stronger associations. In our population-based study, we examined the whole spectrum of symptom severity, which might dilute associations.

Moreover, while we sampled children and adolescents, prior literature generally focused on *adults*. It is possible that more substantial differences in FNC are observed later in life. Yet, some of the largest lifetime changes in brain neuroanatomy occur in childhood (Bethlehem et al., 2022). Further studies in early life are, therefore, necessary. However, the time window we sampled may have several drawbacks as adolescence is characterized by substantial interindividual variation (Galván, 2017; Rutter & Sroufe, 2000).

Inconsistencies with prior literature may also stem from the use of *cross-sectional vs. longitudinal designs*. It is not surprising that a longitudinal design would lead to the identification of smaller associations than a cross-sectional one. Cross-sectional studies, especially those focused on adults, might detect relationships between functional connectivity and psychiatric problems which could reflect life-long brain-behavior relationships. Longitudinal studies can instead dissect associations at a specific time point.

Further, our negative results should be considered in the context of key *challenges in the psychiatric neuroimaging field*: small sample sizes, publication bias, and analytical flexibility. Specifically, small samples are more prone to population variability and thus to capture effects that are inflated by chance (Marek et al., 2022). This problem is reduced as samples get larger (Marek et al., 2022). This may have made our sample less prone to chance findings. Moreover, inflated effects (e.g., due to chance or biases) are more likely to be statistically significant. These findings may be inadvertently overrepresented in the literature (Button et al., 2013; Marek et al., 2022), while negative results may have been overlooked (Button et al., 2013) (i.e., publication bias may have occurred). Such a focus on statistical significance may also make certain analytical choices more preferrable than others (Gelman & Loken, 2013). Overall, when accounting for these key challenges in the field, our negative results may not be that unexpected.

### Strengths and limitations of the present study

Several strengths of this study should be discussed. First, we leveraged a large single-site pediatric sample with repeated measurements on both static and dynamic FNC. Second, we used longitudinal modeling to examine the relationship between FNC and psychiatric problems over time, accounting for changes from childhood to adolescence. This allows to move closer to a causal understanding of these associations by removing prior brain-behavior relationships and disentangling temporal directionality. Third, children were sampled during a key developmental period, in which substantial neuro- and psychiatric development changes occur: brain reorganization takes place and psychiatric problems such as anxiety and depression generally arise or exacerbate (Lee et al., 2014; Vijayakumar et al., 2018).

This study also presents several limitations. We did not leverage an independent sample to test the replicability and generalizability of our findings. Future work is thus required to appropriately evaluate our results. Moreover, we could not account for intraindividual variability across time due to our limited number of repeated assessments. Given the high interindividual differences in FNC, future studies with more than two repeated assessments could explore how relationships between FNC and psychiatric symptoms vary across individuals, both in terms of starting levels (random intercepts) and developmental rates (random slopes). Additionally, resting-state scans in the Generation R sample are relatively short compared to other studies. While their duration is sufficient to reliably capture resting-state patterns in youth (Muetzel et al., 2016; White et al., 2014), there are advantages in adopting a longer scan duration.

## CONCLUSION

We explored the longitudinal relationship between static and dynamic FNC with psychiatric symptoms in a large (*N* = 3,131) population-based sample of youths, leveraging repeated measurements that allowed us to model change over time. Only small, longitudinal relationships were observed. While some associations stood out due to their larger effect sizes, namely for specific psychiatric problems in childhood (i.e., rule-breaking, somatic complaints, thought problems, and attention problems) with dFNC changes into adolescence, these warrant careful interpretations and replication. Future studies should replicate our findings, leverage larger samples, or more repeated assessments to detect smaller effects and model interindividual variation.

## Data Availability

Data for the Generation R Study is available upon request to Vincent Jaddoe.

## Abbreviations

FNC: functional network connectivity
dFNC: dynamic functional network connectivity
sFNC: static functional network connectivity

## KEY POINTS AND RELEVANCE

- Functional network connectivity (FNC) has been proposed as an etiological mechanism to psychiatric problem development, but studies show inconsistencies and are generally small-sized, cross-sectional, and in clinical populations
- This is the first large, longitudinal study of youth from the general population investigating static and dynamic FNC with psychiatric symptoms
- Robust evidence for associations between FNC and psychiatric symptoms was not found
- Specific psychiatric symptoms nominally predicted changes in dynamic FNC but these findings require replication
- Longitudinal designs are key to dissecting when associations occur during development and large samples are needed to have the power to detect the small associations between FNC and psychiatric problems

## ACKNOWLEDGMENTS

Based on the CRediT role taxonomy (https://credit.niso.org/), conceptualization (LDA, FEL, MLV, HT, RM), data curation (LDA, FEL, MLV, BX, OA, EB, KL, VDC, and RM), formal analysis (LDA), funding acquisition (HT, RM), investigation (LDA), methodology (LDA, FEL, MLV, HT, and RM), project administration (LDA, HT, and RM), resources (RM), software (LDA, FEL, MLV, OA, EB, KL, VDC), supervision (HT, RM), validation (FEL and BX), visualization (LDA), writing - original draft (LDA), writing - review & editing (LDA, FEL, MLV, BX OA, EB, KL, VDC, HT, and RM). We thank the participants, general practitioners, hospitals, midwives, and pharmacies in Rotterdam who contributed to the study.

