## Supplementary Materials for "Exploring the longitudinal associations of functional network connectivity and psychiatric symptom changes in youth"

**Table S1.** Descriptive statistics of the sample population

| **Variable** | **Level** | **Value** |
| --- | --- | --- |
| Characteristic path length T1 (mean ± SD) |  | 3.29 ± 0.32 |
| Characteristic path length T2 (mean ± SD) |  | 3.13 ± 0.36 |
| Global efficiency T1 (median, (IQR)) |  | 0.36 (0.34 - 0.39) |
| Global efficiency T2 (median, (IQR)) |  | 0.38 (0.35 - 0.42) |
| Modularity T1 (mean ± SD) |  | 0.16 ± 0.03 |
| Modularity T2 (mean ± SD) |  | 0.16 ± 0.03 |
| Clustering coefficient T1 (median, (IQR)) |  | 0.21 (0.20 - 0.24) |
| Clustering coefficient T2 (median, (IQR)) |  | 0.23 (0.20 - 0.27) |
| MDT state 1 T1 (median, (IQR)) |  | 0 (0 - 1) |
| MDT state 1 T2 (median, (IQR)) |  | 11 (0 - 31) |
| MDT state 2 T1 (median, (IQR)) |  | 12 (5 - 21) |
| MDT state 2 T2 (median, (IQR)) |  | 14 (3- 23) |
| MDT state 3 T1 (median, (IQR)) |  | 14 (8 - 21) |
| MDT state 3 T2 (median, (IQR)) |  | 10 (4 - 18) |
| MDT state 4 T1 (median, (IQR)) |  | 12 (0 - 21) |
| MDT state 4 T2 (median, (IQR)) |  | 10 (0 - 20) |
| MDT state 5 T1 (median, (IQR)) |  | 17 (11 - 25) |
| MDT state 5 T2 (median, (IQR)) |  | 12 (3, 17) |
| NT T1 (mean ± SD) |  | 8.19 ± 2.87 |
| NT T2 (mean ± SD) |  | 7.50 ± 3.32 |
| Internalising problems T1 (median, (IQR)) |  | 1.73 (1.00 - 2.65) |
| Internalising problems T2 (median, (IQR)) |  | 2.03 (1.00 - 2.83) |
| Externalising problems T1 (median, (IQR)) |  | 1.41 (0.00 - 2.24) |
| Externalising problems T2 (median, (IQR)) |  | 1.41 (0.00 - 2.45) |
| Age at MRI assessment T1, years (mean ± SD) |  | 10.12 ± 0.60 |
| Age at MRI assessment T2, years (mean ± SD) |  | 13.95 ± 0.57 |
| Sex (N (%)) | Female | 1,634 (52%) |
|  | Male | 1,496 (48%) |
| Ethnicity/National Origin (N (%)) | Dutch | 1,975 (64%) |
|  | European | 469 (15%) |
|  | Other | 648 (21%) |
| Highest parental education (N (%)) | Low | 67 (2.4%) |
|  | Intermediate | 990 (35%) |
|  | High | 1,748 (62%) |
| Perceived pubertal stage (mean ± SD) |  | 2.43 ± 0.75 |

*Note.* IQR = interquartile range; MDT = mean dwell time; N = number; NT = number of transitions; SD = standard deviation; T = time point

**Table S2.** Non-response analyses comparing the subset with the full sample at T1.

|  | **Summary statistics of each sample** | | | **Comparison across samples** | | |
| --- | --- | --- | --- | --- | --- | --- |
|  |  | Full sample^1^  (*n* = 3,601) | Sub-sample^1^  (*n* = 2,172) | *t* or *X* statistic | df | *p*-value |
| Puberty |  | 2.40 (0.75) | 2.43 (0.75) | 1,055 | 4555,917 | 0.292 |
| Sex | Female | 1,848 (51.3%) | 1,151 (53%) | 1,454 | 1 | 0.228 |
| National origin | Dutch | 2,508 (69.6%) | 1,491 (68.6%) | 0,681 | 2 | 0.711 |
|  | European | 487 (13.5%) | 381 (17.5%) |  |  |  |
|  | Other | 606 (16.8%) | 300 (13.8%) |  |  |  |
| Highest parental education | Low | 78 (2.2%) | 42 (1.9%) | 0,360 | 2 | 0.835 |
|  | Intermediate | 1,180 (32.8%) | 713 (32.8%) |  |  |  |
|  | High | 2,343 (65.1%) | 1,417 (65.2%) |  |  |  |

^1^Sample size after excluding missing values for chosen covariates. For continuous variables, mean and SD are shown. For categorical variables the number and percentage of observations are shown. *Note.* Df = degrees of freedom; SD = standard deviation

**Table S3.** Model fits for the cross-lagged panel models for static functional network connectivity and internalizing and externalizing problems.

|  | **RMSEA** | **SRMR** | **CFI** | **TLI** |
| --- | --- | --- | --- | --- |
| *Internalizing problems* |  |  |  |  |
| Characteristic path length | 0.023 | 0.020 | 0.960 | 0.938 |
| Global efficiency | 0.021 | 0.019 | 0.965 | 0.945 |
| Modularity | 0.031 | 0.027 | 0.926 | 0.886 |
| Clustering coefficient | 0.021 | 0.019 | 0.964 | 0.944 |
| *Externalizing problems* |  |  |  |  |
| Characteristic path length | 0.029 | 0.02 | 0.945 | 0.914 |
| Global efficiency | 0.028 | 0.019 | 0.948 | 0.920 |
| Modularity | 0.036 | 0.026 | 0.916 | 0.869 |
| Clustering coefficient | 0.028 | 0.019 | 0.947 | 0.918 |

*Note.* CFI = comparative fit index; RMSEA = root mean square error of approximation; SRMR = standardized root mean square residual; TLI = Tucker-Lewis index.

**Table S4.** Results for the cross-lagged panel models for static functional network connectivity and internalizing and externalizing problems after stricter motion quality control

| **Model** | **Standardized estimate** | **SE** | ***p*-value** |
| --- | --- | --- | --- |
| *Static FNC T1 🡪 Psychiatric symptoms T2 (corrected for T1 symptoms)* |  |  |  |
| Internalizing problems T2 ~ Characteristic path length T1 | 0.025 | 0.019 | 0.200 |
| Internalizing problems T2 ~ Global efficiency T1 | -0.023 | 0.019 | 0.233 |
| Internalizing problems T2 ~ Modularity T1 | -0.014 | 0.019 | 0.457 |
| Internalizing problems T2 ~ Clustering coefficient T1 | -0.016 | 0.019 | 0.387 |
| Externalizing problems T2 ~ Characteristic path length T1 | 0.020 | 0.019 | 0.294 |
| Externalizing problems T2 ~ Global efficiency T1 | -0.022 | 0.019 | 0.229 |
| Externalizing problems T2 ~ Modularity T1 | 0.004 | 0.019 | 0.850 |
| Externalizing problems T2 ~ Clustering coefficient T1 | -0.022 | 0.019 | 0.233 |
| *Psychiatric symptoms T1 🡪 Static FNC T2 (corrected for T1 FNC)* |  |  |  |
| Characteristic path length T2 ~ Internalizing problems T1 | 0.03 | 0.026 | 0.244 |
| Global efficiency T2 ~ Internalizing problems T1 | -0.032 | 0.026 | 0.218 |
| Modularity T2 ~ Internalizing problems T1 | -0.02 | 0.026 | 0.44 |
| Clustering coefficient T2 ~ Internalizing problems T1 | -0.026 | 0.026 | 0.331 |
| Characteristic path length T2 ~ Externalizing problems T1 | -0.008 | 0.026 | 0.763 |
| Global efficiency T2 ~ Externalizing problems T1 | 0.007 | 0.027 | 0.791 |
| Modularity T2 ~ Externalizing problems T1 | -0.033 | 0.026 | 0.214 |
| Clustering coefficient T2 ~ Externalizing problems T1 | 0.013 | 0.027 | 0.637 |

*Note.* ~ = regressed on; SE = standard error; T = time point

**Table S5.** Differences across sexes for static functional network connectivity with internalizing and externalizing problems.

|  | **Df** | **AIC** | **BIC** | **Chisq** | **Chisq diff** | **Df diff** | ***p*-value** |
| --- | --- | --- | --- | --- | --- | --- | --- |
| *Internalizing problems* |  |  |  |  |  |  |  |
| Characteristic path length (base) | 48 | 62641.806 | 63440.246 | 83.369 | NA | NA | NA |
| Characteristic path length (equal reg.) | 60 | 62623.899 | 63349.754 | 89.462 | 6.093 | 12 | 0.911 |
| Global efficiency (base) | 48 | 47075.635 | 47874.075 | 77.128 | NA | NA | NA |
| Global efficiency (equal reg.) | 60 | 47058.183 | 47784.038 | 83.676 | 6.548 | 12 | 0.886 |
| Modularity (base) | 48 | 42617.105 | 43415.545 | 85.088 | NA | NA | NA |
| Modularity (equal reg.) | 60 | 42600.072 | 43325.927 | 92.055 | 6.967 | 12 | 0.860 |
| Clustering coefficient (base) | 48 | 46883.834 | 47682.274 | 78.491 | NA | NA | NA |
| Clustering coefficient (equal reg.) | 60 | 46865.159 | 47591.014 | 83.816 | 5.325 | 12 | 0.946 |
| *Externalizing problems* |  |  |  |  |  |  |  |
| Characteristic path length (base) | 48 | 62837.594 | 63636.034 | 93.255 | NA | NA | NA |
| Characteristic path length (equal reg.) | 60 | 62821.405 | 63547.260 | 101.067 | 7.812 | 12 | 0.800 |
| Global efficiency (base) | 48 | 47271.817 | 48070.257 | 87.146 | NA | NA | NA |
| Global efficiency (equal reg.) | 60 | 47256.446 | 47982.301 | 95.775 | 8.629 | 12 | 0.734 |
| Modularity (base) | 48 | 42809.782 | 43608.222 | 95.201 | NA | NA | NA |
| Modularity (equal reg.) | 60 | 42793.243 | 43519.098 | 102.662 | 7.462 | 12 | 0.826 |
| Clustering coefficient (base) | 48 | 47080.563 | 47879.003 | 88.767 | NA | NA | NA |
| Clustering coefficient (equal reg.) | 60 | 47063.403 | 47789.257 | 95.606 | 6.839 | 12 | 0.868 |

*Note.* AIC = Akaike’s Information Criterio; BIC = Bayesian Information Criterion; Chisq = chi-square; Diff = difference; df = degrees of freedom; NA = not applicable; reg = regression coefficients

**Table S6.** Longitudinal relations between static functional network connectivity and specific psychiatric problems from late childhood to early adolescence.

| **Model** | **Standardized estimate** | **SE** | ***p*-value** |
| --- | --- | --- | --- |
| *Static FNC T1 🡪 Psychiatric symptoms T2 (corrected for T1 symptoms)* |  |  |  |
| Anxious/Depressed T2 ~ Modularity T1 | -0.025 | 0.020 | 0.207 |
| Withdrawn/Depressed T2 ~ Modularity T1 | -0.033 | 0.021 | 0.109 |
| Somatic complaints T2 ~ Modularity T1 | -0.009 | 0.021 | 0.672 |
| Social problems T2 ~ Modularity T1 | -0.013 | 0.020 | 0.518 |
| Thought problems T2 ~ Modularity T1 | 0.009 | 0.020 | 0.652 |
| Attention problems T2 ~ Modularity T1 | 0.016 | 0.019 | 0.397 |
| Aggressive behaviors T2 ~ Modularity T1 | -0.002 | 0.018 | 0.916 |
| Rule-breaking behaviors T2 ~ Modularity T1 | 0.012 | 0.021 | 0.564 |
| Anxious/Depressed T2 ~ Clustering coefficient T1 | -0.008 | 0.020 | 0.692 |
| Withdrawn/Depressed T2 ~ Clustering coefficient T1 | -0.005 | 0.020 | 0.796 |
| Somatic complaints T2 ~ Clustering coefficient T1 | 0.002 | 0.021 | 0.923 |
| Social problems T2 ~ Clustering coefficient T1 | -0.005 | 0.020 | 0.801 |
| Thought problems T2 ~ Clustering coefficient T1 | -0.010 | 0.020 | 0.629 |
| Attention problems T2 ~ Clustering coefficient T1 | -0.016 | 0.019 | 0.403 |
| Aggressive behaviors T2 ~ Clustering coefficient T1 | -0.012 | 0.018 | 0.512 |
| Rule-breaking behaviors T2 ~ Clustering coefficient T1 | -0.010 | 0.021 | 0.637 |
| Anxious/Depressed T2 ~ Characteristic path length T1 | 0.016 | 0.020 | 0.434 |
| Withdrawn/Depressed T2 ~ Characteristic path length T1 | 0.015 | 0.021 | 0.477 |
| Somatic complaints T2 ~ Characteristic path length T1 | 0.003 | 0.021 | 0.885 |
| Social problems T2 ~ Characteristic path length T1 | 0.006 | 0.020 | 0.768 |
| Thought problems T2 ~ Characteristic path length T1 | 0.012 | 0.020 | 0.551 |
| Attention problems T2 ~ Characteristic path length T1 | 0.013 | 0.019 | 0.489 |
| Aggressive behaviors T2 ~ Characteristic path length T1 | 0.010 | 0.018 | 0.599 |
| Rule-breaking behaviors T2 ~ Characteristic path length T1 | 0.012 | 0.021 | 0.570 |
| Anxious/Depressed T2 ~ Global efficiency T1 | -0.014 | 0.020 | 0.479 |
| Withdrawn/Depressed T2 ~ Global efficiency T1 | -0.011 | 0.020 | 0.579 |
| Somatic complaints T2 ~ Global efficiency T1 | -0.001 | 0.021 | 0.976 |
| Social problems T2 ~ Global efficiency T1 | -0.007 | 0.020 | 0.718 |
| Thought problems T2 ~ Global efficiency T1 | -0.012 | 0.020 | 0.539 |
| Attention problems T2 ~ Global efficiency T1 | -0.012 | 0.019 | 0.517 |
| Aggressive behaviors T2 ~ Global efficiency T1 | -0.013 | 0.018 | 0.469 |
| Rule-breaking behaviors T2 ~ Global efficiency T1 | -0.007 | 0.021 | 0.729 |
| *Psychiatric symptoms T1 🡪 Static FNC T2 (corrected for T1 FNC)* |  |  |  |
| Modularity T2 ~ Anxious/Depressed T1 | -0.020 | 0.025 | 0.427 |
| Modularity T2 ~ Withdrawn/Depressed T1 | 0.013 | 0.025 | 0.599 |
| Modularity T2 ~ Somatic complaints T1 | -0.042 | 0.025 | 0.088 |
| Modularity T2 ~ Social problems T1 | -0.021 | 0.024 | 0.386 |
| Modularity T2 ~ Thought problems T1 | -0.018 | 0.024 | 0.473 |
| Modularity T2 ~ Attention problems T1 | -0.016 | 0.024 | 0.513 |
| Modularity T2 ~ Aggressive behaviors T1 | -0.026 | 0.024 | 0.288 |
| Modularity T2 ~ Rule-breaking behaviors T1 | -0.047 | 0.025 | 0.056 |
| Clustering coefficient T2 ~ Anxious/Depressed T1 | -0.034 | 0.025 | 0.170 |
| Clustering coefficient T2 ~ Withdrawn/Depressed T1 | -0.021 | 0.025 | 0.401 |
| Clustering coefficient T2 ~ Somatic complaints T1 | 0.018 | 0.025 | 0.474 |
| Clustering coefficient T2 ~ Social problems T1 | -0.005 | 0.024 | 0.844 |
| Clustering coefficient T2 ~ Thought problems T1 | -0.006 | 0.025 | 0.821 |
| Clustering coefficient T2 ~ Attention problems T1 | -0.019 | 0.024 | 0.423 |
| Clustering coefficient T2 ~ Aggressive behaviors T1 | 0.007 | 0.025 | 0.790 |
| Clustering coefficient T2 ~ Rule-breaking behaviors T1 | 0.026 | 0.025 | 0.292 |
| Characteristic path length T2 ~ Anxious/Depressed T1 | 0.038 | 0.025 | 0.123 |
| Characteristic path length T2 ~ Withdrawn/Depressed T1 | 0.027 | 0.024 | 0.265 |
| Characteristic path length T2 ~ Somatic complaints T1 | -0.007 | 0.025 | 0.780 |
| Characteristic path length T2 ~ Social problems T1 | 0.007 | 0.024 | 0.767 |
| Characteristic path length T2 ~ Thought problems T1 | 0.011 | 0.024 | 0.643 |
| Characteristic path length T2 ~ Attention problems T1 | 0.021 | 0.024 | 0.378 |
| Characteristic path length T2 ~ Aggressive behaviors T1 | -0.008 | 0.024 | 0.748 |
| Characteristic path length T2 ~ Rule-breaking behaviors T1 | -0.018 | 0.025 | 0.457 |
| Global efficiency T2 ~ Anxious/Depressed T1 | -0.042 | 0.025 | 0.086 |
| Global efficiency T2 ~ Withdrawn/Depressed T1 | -0.023 | 0.025 | 0.343 |
| Global efficiency T2 ~ Somatic complaints T1 | 0.010 | 0.025 | 0.695 |
| Global efficiency T2 ~ Social problems T1 | -0.012 | 0.024 | 0.623 |
| Global efficiency T2 ~ Thought problems T1 | -0.013 | 0.024 | 0.582 |
| Global efficiency T2 ~ Attention problems T1 | -0.029 | 0.024 | 0.233 |
| Global efficiency T2 ~ Aggressive behaviors T1 | 0.003 | 0.025 | 0.913 |
| Global efficiency T2 ~ Rule-breaking behaviors T1 | 0.021 | 0.025 | 0.392 |

*Note.* ~ = regressed on; SE = standard error; T = time point.

**Table S7.** Model fits for the cross-lagged panel models for dynamic functional network connectivity with internalizing and externalizing problems.

|  | **RMSEA** | **SRMR** | **CFI** | **TLI** |
| --- | --- | --- | --- | --- |
| *Internalizing problems* | | | | |
| MDT1 | 0.030 | 0.024 | 0.932 | 0.894 |
| MDT2 | 0.023 | 0.020 | 0.957 | 0.934 |
| MDT3 | 0.022 | 0.018 | 0.963 | 0.942 |
| MDT4 | 0.023 | 0.020 | 0.960 | 0.937 |
| MDT5 | 0.020 | 0.019 | 0.967 | 0.948 |
| NT | 0.018 | 0.018 | 0.974 | 0.959 |
| *Externalizing problems* | |  |  |  |
| MDT1 | 0.035 | 0.024 | 0.921 | 0.877 |
| MDT2 | 0.029 | 0.020 | 0.943 | 0.912 |
| MDT3 | 0.028 | 0.018 | 0.946 | 0.916 |
| MDT4 | 0.030 | 0.020 | 0.944 | 0.912 |
| MDT5 | 0.028 | 0.019 | 0.949 | 0.921 |
| NT | 0.026 | 0.018 | 0.955 | 0.929 |

*Note.* CFI = comparative fit index; MDT = mean dwell time; NT = number of transitions; RMSEA = root mean square error of approximation; SRMR = standardized root mean square residual; TLI = Tucker-Lewis index.

**Table S8.** Results for the cross-lagged panel models for dynamic functional network connectivity and internalizing and externalizing problems after stricter motion quality control

| **Model** | **Standardized estimate** | **SE** | **p-value** |
| --- | --- | --- | --- |
| *Dynamic FNC T1 🡪 Psychiatric symptoms T2 (corrected for T1 symptoms)* | | | |
| Internalizing problems T2 ~ MDT state 1 T1 | 0.007 | 0.019 | 0.717 |
| Internalizing problems T2 ~ MDT state 2 T1 | -0.014 | 0.019 | 0.469 |
| Internalizing problems T2 ~ MDT state 3 T1 | 0.003 | 0.019 | 0.862 |
| Internalizing problems T2 ~ MDT state 4 T1 | 0.028 | 0.019 | 0.143 |
| Internalizing problems T2 ~ MDT state 5 T1 | -0.015 | 0.019 | 0.416 |
| Internalizing problems T2 ~ NT T1 | 0.009 | 0.019 | 0.625 |
| Externalizing problems T2 ~ MDT state 1 T1 | 0.020 | 0.019 | 0.295 |
| Externalizing problems T2 ~ MDT state 2 T1 | 0.017 | 0.018 | 0.369 |
| Externalizing problems T2 ~ MDT state 3 T1 | 0.025 | 0.019 | 0.182 |
| Externalizing problems T2 ~ MDT state 4 T1 | 0.025 | 0.019 | 0.189 |
| Externalizing problems T2 ~ MDT state 5 T1 | -0.048 | 0.019 | 0.010 |
| Externalizing problems T2 ~ NT T1 | 0.018 | 0.019 | 0.349 |
| *Psychiatric symptoms T1 🡪 Dynamic FNC T2 (corrected for T1 FNC)* | | | |
| MDT state 1 T2 ~ Internalizing problems T1 | -0.027 | 0.027 | 0.315 |
| MDT state 2 T2 ~ Internalizing problems T1 | -0.004 | 0.026 | 0.870 |
| MDT state 3 T2 ~ Internalizing problems T1 | -0.040 | 0.026 | 0.121 |
| MDT state 4 T2 ~ Internalizing problems T1 | 0.008 | 0.025 | 0.758 |
| MDT state 5 T2 ~ Internalizing problems T1 | 0.031 | 0.027 | 0.248 |
| NT T2 ~ Internalizing problems T1 | 0.022 | 0.026 | 0.395 |
| MDT state 1 T2 ~ Externalizing problems T1 | 0.026 | 0.027 | 0.343 |
| MDT state 2 T2 ~ Externalizing problems T1 | -0.036 | 0.026 | 0.166 |
| MDT state 3 T2 ~ Externalizing problems T1 | -0.038 | 0.026 | 0.145 |
| MDT state 4 T2 ~ Externalizing problems T1 | 0.006 | 0.026 | 0.809 |
| MDT state 5 T2 ~ Externalizing problems T1 | 0.025 | 0.027 | 0.363 |
| NT T2 ~ Externalizing problems T1 | 0.012 | 0.027 | 0.659 |

*Note.* ~ = regressed on; MDT = mean dwell time; NT = number of transitions; T = time-point

**Table S9.** Comparison across sexes for relations of dynamic functional network connectivity and internalizing and externalizing problems.

|  | **Df** | **AIC** | **BIC** | **Chisq** | **Chisq diff** | **Df diff** | ***p-*value** |
| --- | --- | --- | --- | --- | --- | --- | --- |
| *Internalizing problems* | | | | | | | |
| MDT1 (base) | 48 | 39513.422 | 40311.862 | 130.948 | NA | NA | NA |
| MDT1 (equal reg) | 60 | 39494.843 | 40220.698 | 136.369 | 5.421 | 12 | 0.942 |
| MDT2 (base) | 48 | 37820.222 | 38618.662 | 64.335 | NA | NA | NA |
| MDT2 (equal reg) | 60 | 37810.188 | 38536.043 | 78.300 | 13.966 | 12 | 0.303 |
| MDT3 (base) | 48 | 35426.176 | 36224.616 | 79.027 | NA | NA | NA |
| MDT3 (equal reg) | 60 | 35416.027 | 36141.881 | 92.878 | 13.851 | 12 | 0.310 |
| MDT4 (base) | 48 | 38457.106 | 39255.546 | 64.752 | NA | NA | NA |
| MDT4 (equal reg) | 60 | 38446.574 | 39172.429 | 78.221 | 13.468 | 12 | 0.336 |
| MDT5 (base) | 48 | 35800.693 | 36599.133 | 74.023 | NA | NA | NA |
| MDT5 (equal reg) | 60 | 35782.078 | 36507.932 | 79.407 | 5.384 | 12 | 0.944 |
| NT (base) | 48 | 61897.294 | 62695.734 | 60.479 | NA | NA | NA |
| NT (equal reg) | 60 | 61885.199 | 62611.054 | 72.384 | 11.905 | 12 | 0.453 |
| *Externalizing problems* | | | | | | | |
| MDT1 (base) | 48 | 39703.432 | 40501.872 | 140.794 | NA | NA | NA |
| MDT1 (equal reg) | 60 | 39686.968 | 40412.823 | 148.33 | 7.536 | 12 | 0.820 |
| MDT2 (base) | 48 | 38008.752 | 38807.192 | 75.939 | NA | NA | NA |
| MDT2 (equal reg) | 60 | 38001.254 | 38727.109 | 92.441 | 16.502 | 12 | 0.169 |
| MDT3 (base) | 48 | 35625.368 | 36423.808 | 90.833 | NA | NA | NA |
| MDT3 (equal reg) | 60 | 35612.247 | 36338.101 | 101.711 | 10.878 | 12 | 0.539 |
| MDT4 (base) | 48 | 38651.600 | 39450.040 | 74.445 | NA | NA | NA |
| MDT4 (equal reg) | 60 | 38643.909 | 39369.764 | 90.754 | 16.309 | 12 | 0.178 |
| MDT5 (base) | 48 | 35987.356 | 36785.796 | 84.311 | NA | NA | NA |
| MDT5 (equal reg) | 60 | 35969.902 | 36695.756 | 90.857 | 6.546 | 12 | 0.886 |
| NT (base) | 48 | 62100.107 | 62898.547 | 71.441 | NA | NA | NA |
| NT (equal reg) | 60 | 62086.902 | 62812.757 | 82.237 | 10.796 | 12 | 0.546 |

*Note.* AIC = Akaike’s Information Criterion; BIC = Bayesian Information Criterion; Chisq = chi-square; Diff = difference; df = degrees of freedom; ext = externalizing; int = internalizing; MDT = mean dwell time; NA = not applicable; nt = number of transitions;

reg = regression coefficients

**Table S10.** Longitudinal relationships between dynamic functional network connectivity and specific psychiatric problems from late childhood to early adolescence

| **Model** | **Standardized estimate** | **SE** | ***p*-value** |
| --- | --- | --- | --- |
| *Dynamic FNC T1 🡪 Psychiatric symptoms T2 (corrected for T1 symptoms)* | | | |
| Anxious/Depressed T2 ~ MDT state 1 T1 | 0.007 | 0.020 | 0.725 |
| Withdrawn/Depressed T2 ~ MDT state 1 T1 | 0.010 | 0.021 | 0.641 |
| Somatic complaints T2 ~ MDT state 1 T1 | 0.023 | 0.022 | 0.281 |
| Social problems T2 ~ MDT state 1 T1 | 0.034 | 0.021 | 0.096 |
| Thought problems T2 ~ MDT state 1 T1 | 0.024 | 0.021 | 0.254 |
| Attention problems T2 ~ MDT state 1 T1 | -0.001 | 0.019 | 0.940 |
| Aggressive behaviors T2 ~ MDT state 1 T1 | 0.025 | 0.019 | 0.184 |
| Rule-breaking behaviors T2 ~ MDT state 1 T1 | 0.030 | 0.021 | 0.165 |
| Anxious/Depressed T2 ~ MDT state 2 T1 | 0.002 | 0.020 | 0.932 |
| Withdrawn/Depressed T2 ~ MDT state 2 T1 | -0.004 | 0.020 | 0.837 |
| Somatic complaints T2 ~ MDT state 2 T1 | -0.016 | 0.021 | 0.440 |
| Social problems T2 ~ MDT state 2 T1 | -0.014 | 0.020 | 0.464 |
| Thought problems T2 ~ MDT state 2 T1 | 0.009 | 0.020 | 0.662 |
| Attention problems T2 ~ MDT state 2 T1 | 0.016 | 0.018 | 0.394 |
| Aggressive behaviors T2 ~ MDT state 2 T1 | 0.013 | 0.018 | 0.472 |
| Rule-breaking behaviors T2 ~ MDT state 2 T1 | 0.007 | 0.021 | 0.737 |
| Anxious/Depressed T2 ~ MDT state 3 T1 | -0.008 | 0.020 | 0.685 |
| Withdrawn/Depressed T2 ~ MDT state 3 T1 | 0.013 | 0.021 | 0.535 |
| Somatic complaints T2 ~ MDT state 3 T1 | 0.014 | 0.021 | 0.516 |
| Social problems T2 ~ MDT state 3 T1 | 0.009 | 0.020 | 0.660 |
| Thought problems T2 ~ MDT state 3 T1 | 0.010 | 0.020 | 0.632 |
| Attention problems T2 ~ MDT state 3 T1 | 0.006 | 0.019 | 0.738 |
| Aggressive behaviors T2 ~ MDT state 3 T1 | 0.016 | 0.018 | 0.389 |
| Rule-breaking behaviors T2 ~ MDT state 3 T1 | 0.041 | 0.021 | 0.052 |
| Anxious/Depressed T2 ~ MDT state 4 T1 | 0.031 | 0.020 | 0.132 |
| Withdrawn/Depressed T2 ~ MDT state 4 T1 | 0.024 | 0.021 | 0.247 |
| Somatic complaints T2 ~ MDT state 4 T1 | 0.026 | 0.021 | 0.223 |
| Social problems T2 ~ MDT state 4 T1 | 0.009 | 0.020 | 0.649 |
| Thought problems T2 ~ MDT state 4 T1 | 0.009 | 0.021 | 0.657 |
| Attention problems T2 ~ MDT state 4 T1 | 0.022 | 0.019 | 0.249 |
| Aggressive behaviors T2 ~ MDT state 4 T1 | 0.02 | 0.019 | 0.294 |
| Rule-breaking behaviors T2 ~ MDT state 4 T1 | 0.019 | 0.021 | 0.370 |
| Anxious/Depressed T2 ~ MDT state 5 T1 | -0.026 | 0.020 | 0.193 |
| Withdrawn/Depressed T2 ~ MDT state 5 T1 | 0.000 | 0.021 | 0.985 |
| Somatic complaints T2 ~ MDT state 5 T1 | -0.005 | 0.021 | 0.814 |
| Social problems T2 ~ MDT state 5 T1 | -0.030 | 0.020 | 0.136 |
| Thought problems T2 ~ MDT state 5 T1 | -0.005 | 0.020 | 0.812 |
| Attention problems T2 ~ MDT state 5 T1 | -0.056 | 0.019 | 0.003 |
| Aggressive behaviors T2 ~ MDT state 5 T1 | -0.056 | 0.018 | 0.002 |
| Rule-breaking behaviors T2 ~ MDT state 5 T1 | -0.017 | 0.021 | 0.406 |
| Anxious/Depressed T2 ~ NT T1 | 0.017 | 0.020 | 0.393 |
| Withdrawn/Depressed T2 ~ NT T1 | -0.006 | 0.021 | 0.779 |
| Somatic complaints T2 ~ NT T1 | -0.018 | 0.021 | 0.400 |
| Social problems T2 ~ NT T1 | 0.031 | 0.020 | 0.118 |
| Thought problems T2 ~ NT T1 | -0.012 | 0.020 | 0.553 |
| Attention problems T2 ~ NT T1 | 0.019 | 0.019 | 0.310 |
| Aggressive behaviors T2 ~ NT T1 | 0.030 | 0.018 | 0.104 |
| Rule-breaking behaviors T2 ~ NT T1 | -0.015 | 0.021 | 0.474 |
| *Psychiatric symptoms T1 🡪 Dynamic FNC T2 (corrected for T1 FNC)* | | | |
| MDT state 1 T2 ~ Anxious/Depressed T1 | -0.042 | 0.025 | 0.089 |
| MDT state 1 T2 ~ Withdrawn/Depressed T1 | -0.033 | 0.025 | 0.180 |
| MDT state 1 T2 ~ Somatic complaints T1 | 0.027 | 0.025 | 0.273 |
| MDT state 1 T2 ~ Social problems T1 | -0.021 | 0.025 | 0.402 |
| MDT state 1 T2 ~ Thought problems T1 | -0.022 | 0.025 | 0.366 |
| MDT state 1 T2 ~ Attention problems T1 | -0.012 | 0.024 | 0.612 |
| MDT state 1 T2 ~ Aggressive behaviors T1 | 0.011 | 0.025 | 0.674 |
| MDT state 1 T2 ~ Rule-breaking behaviors T1 | 0.042 | 0.025 | 0.097 |
| MDT state 2 T2 ~ Anxious/Depressed T1 | 0.031 | 0.024 | 0.197 |
| MDT state 2 T2 ~ Withdrawn/Depressed T1 | 0.001 | 0.024 | 0.975 |
| MDT state 2 T2 ~ Somatic complaints T1 | -0.032 | 0.024 | 0.193 |
| MDT state 2 T2 ~ Social problems T1 | -0.034 | 0.024 | 0.154 |
| MDT state 2 T2 ~ Thought problems T1 | -0.032 | 0.024 | 0.183 |
| MDT state 2 T2 ~ Attention problems T1 | -0.037 | 0.024 | 0.118 |
| MDT state 2 T2 ~ Aggressive behaviors T1 | -0.018 | 0.024 | 0.448 |
| MDT state 2 T2 ~ Rule-breaking behaviors T1 | -0.067 | 0.024 | 0.006 |
| MDT state 3 T2 ~ Anxious/Depressed T1 | -0.042 | 0.025 | 0.086 |
| MDT state 3 T2 ~ Withdrawn/Depressed T1 | -0.027 | 0.025 | 0.265 |
| MDT state 3 T2 ~ Somatic complaints T1 | -0.080 | 0.025 | 0.001 |
| MDT state 3 T2 ~ Social problems T1 | -0.028 | 0.024 | 0.246 |
| MDT state 3 T2 ~ Thought problems T1 | -0.055 | 0.024 | 0.023 |
| MDT state 3 T2 ~ Attention problems T1 | -0.030 | 0.024 | 0.216 |
| MDT state 3 T2 ~ Aggressive behaviors T1 | -0.044 | 0.024 | 0.073 |
| MDT state 3 T2 ~ Rule-breaking behaviors T1 | -0.030 | 0.025 | 0.221 |
| MDT state 4 T2 ~ Anxious/Depressed T1 | -0.015 | 0.024 | 0.526 |
| MDT state 4 T2 ~ Withdrawn/Depressed T1 | -0.021 | 0.024 | 0.380 |
| MDT state 4 T2 ~ Somatic complaints T1 | 0.002 | 0.024 | 0.924 |
| MDT state 4 T2 ~ Social problems T1 | 0.016 | 0.023 | 0.500 |
| MDT state 4 T2 ~ Thought problems T1 | 0.028 | 0.024 | 0.243 |
| MDT state 4 T2 ~ Attention problems T1 | 0.056 | 0.023 | 0.017 |
| MDT state 4 T2 ~ Aggressive behaviors T1 | -0.014 | 0.024 | 0.568 |
| MDT state 4 T2 ~ Rule-breaking behaviors T1 | -0.026 | 0.024 | 0.276 |
| MDT state 5 T2 ~ Anxious/Depressed T1 | 0.024 | 0.025 | 0.340 |
| MDT state 5 T2 ~ Withdrawn/Depressed T1 | 0.033 | 0.025 | 0.193 |
| MDT state 5 T2 ~ Somatic complaints T1 | 0.012 | 0.025 | 0.641 |
| MDT state 5 T2 ~ Social problems T1 | 0.029 | 0.025 | 0.249 |
| MDT state 5 T2 ~ Thought problems T1 | 0.022 | 0.025 | 0.369 |
| MDT state 5 T2 ~ Attention problems T1 | 0.012 | 0.024 | 0.633 |
| MDT state 5 T2 ~ Aggressive behaviors T1 | 0.025 | 0.025 | 0.318 |
| MDT state 5 T2 ~ Rule-breaking behaviors T1 | 0.002 | 0.025 | 0.941 |
| NT T2 ~ Anxious/Depressed T1 | 0.033 | 0.025 | 0.177 |
| NT T2 ~ Withdrawn/Depressed T1 | 0.038 | 0.025 | 0.126 |
| NT T2 ~ Somatic complaints T1 | 0.011 | 0.025 | 0.652 |
| NT T2 ~ Social problems T1 | 0.045 | 0.024 | 0.062 |
| NT T2 ~ Thought problems T1 | 0.067 | 0.024 | 0.006 |
| NT T2 ~ Attention problems T1 | 0.041 | 0.024 | 0.089 |
| NT T2 ~ Aggressive behaviors T1 | 0.020 | 0.025 | 0.408 |
| NT T2 ~ Rule-breaking behaviors T1 | 0.013 | 0.025 | 0.613 |

*Note.* ~ = regressed on; MDT = mean dwell time; NT = number of transitions; SE = standard error; T = time point.

**Figure S1.** Dynamic functional network connectivity states

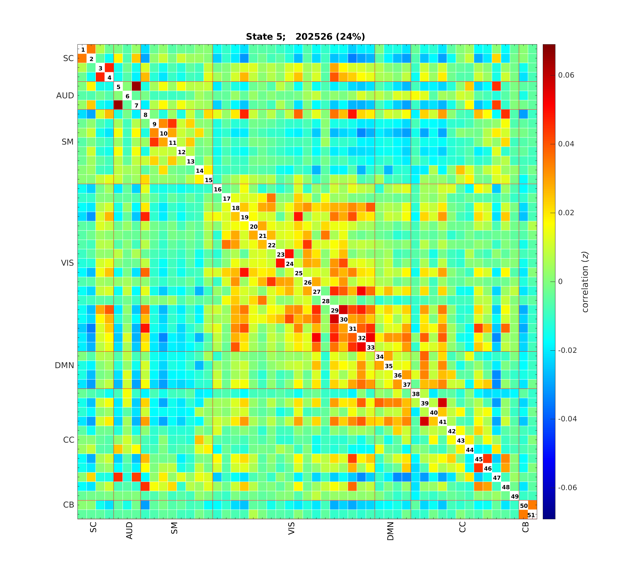

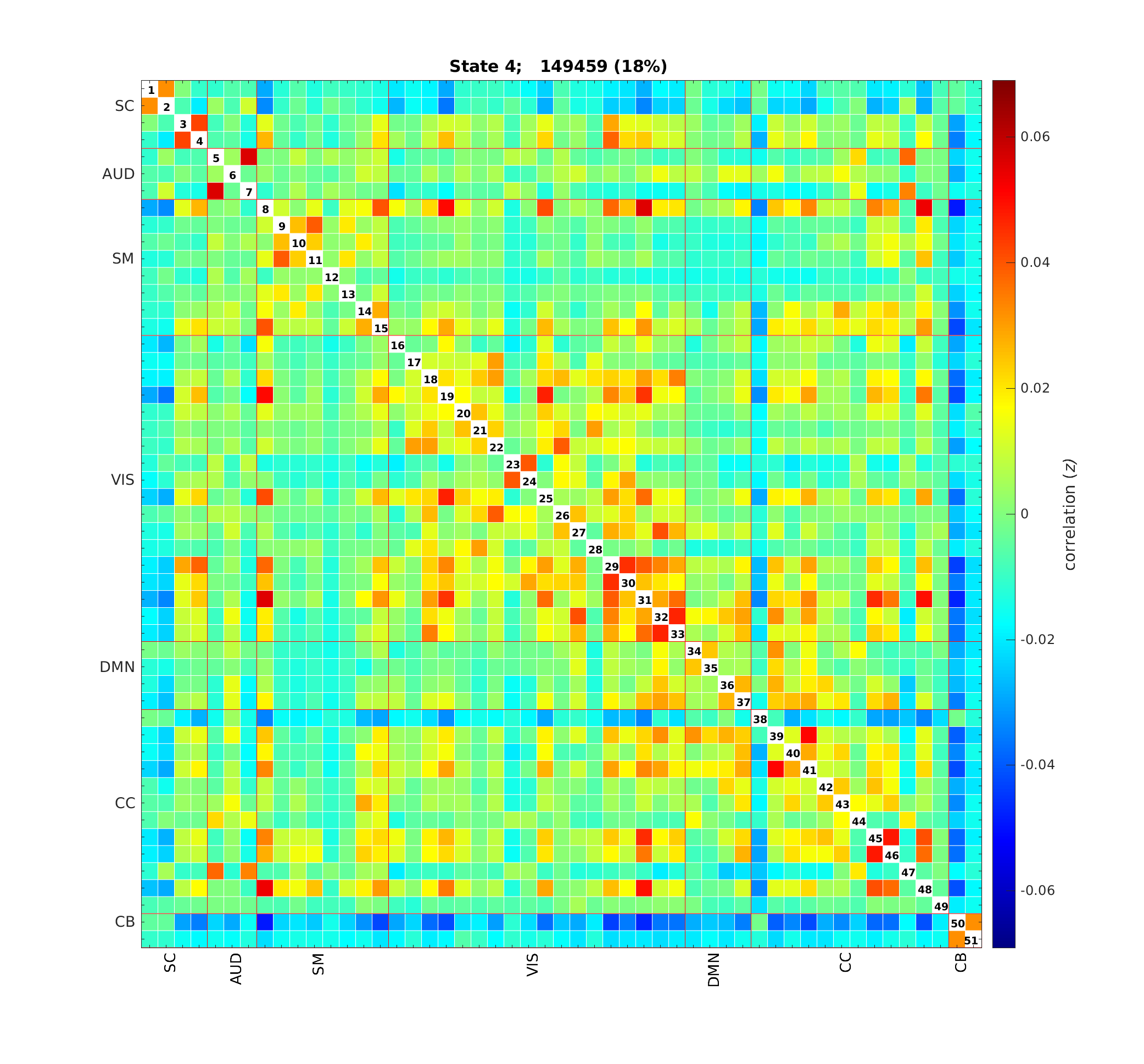

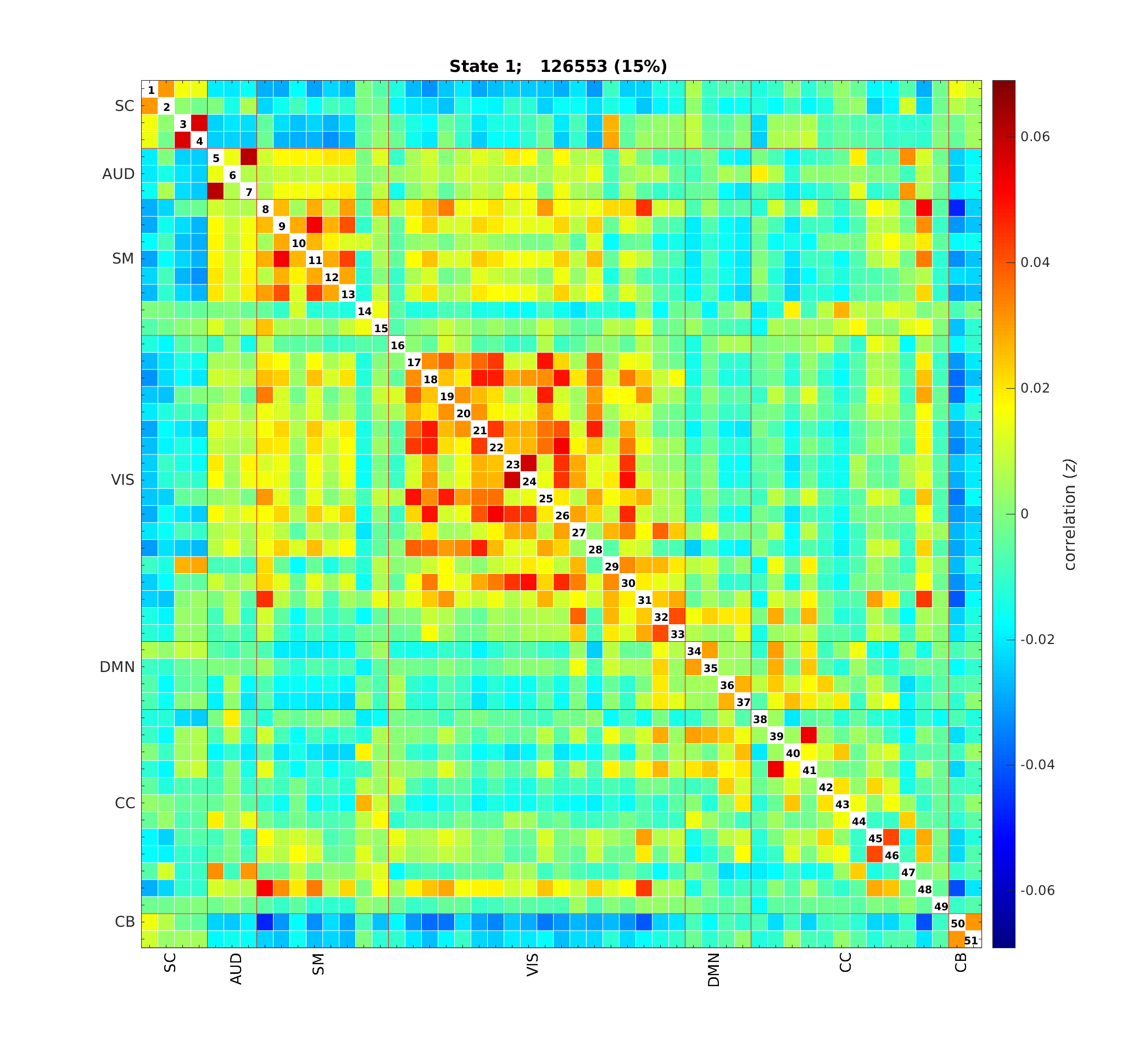

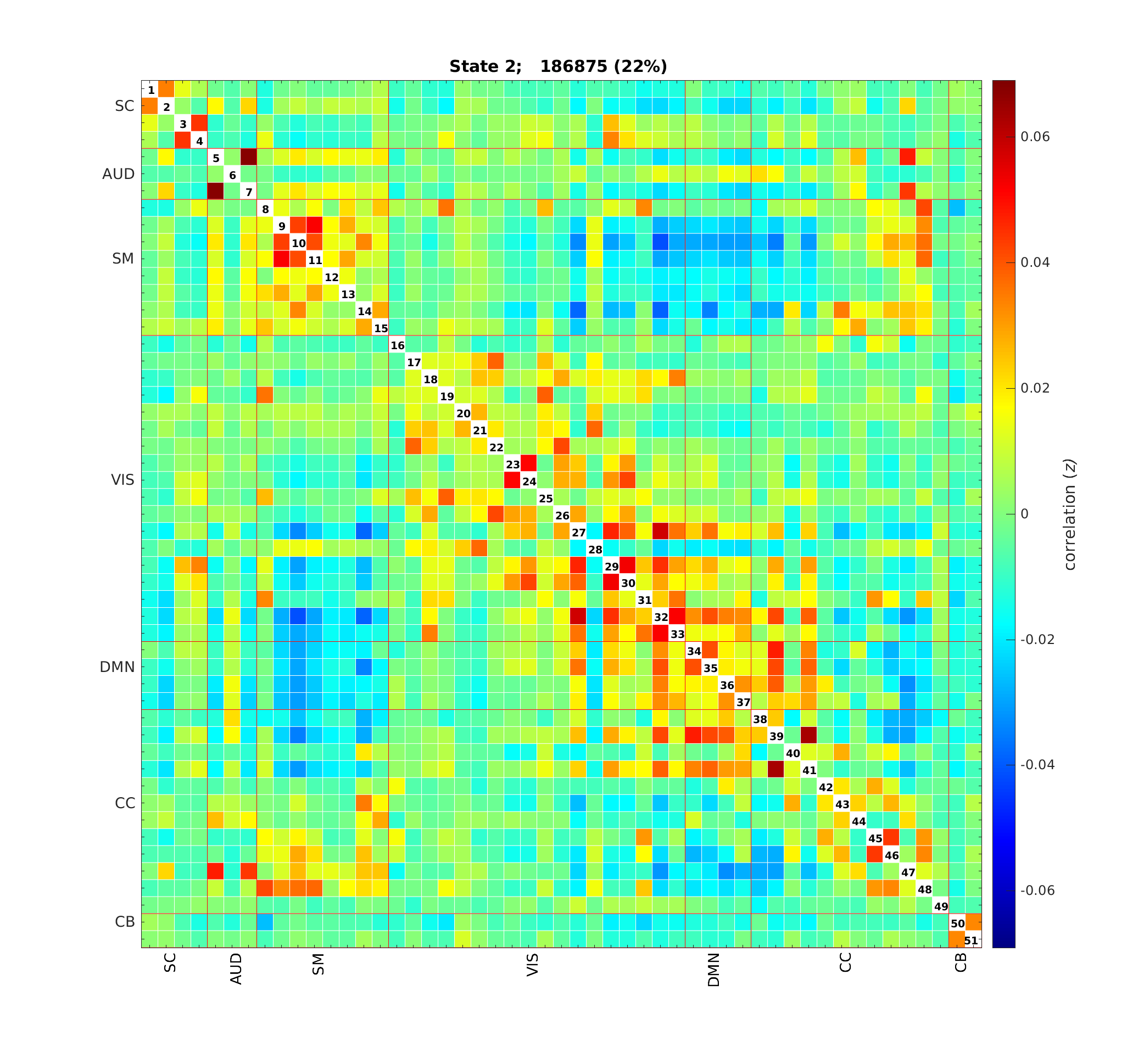

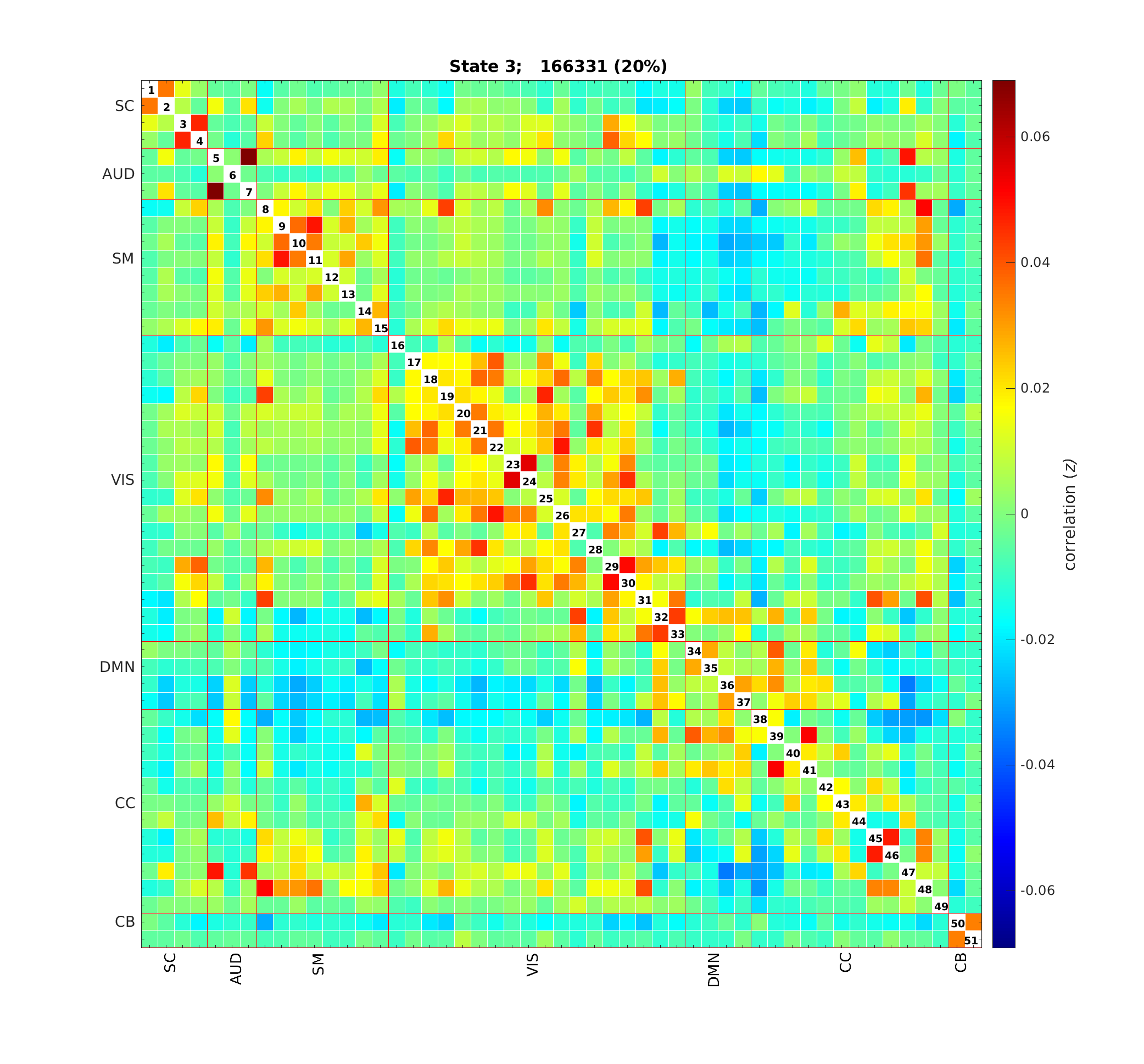

*Note.* The five dynamic functional network connectivity states, which capture the connectivity configurations participants showed during the MRI scanning session. AUD = auditory network; CB = cerebellar network; CC = cognitive control network; DMN = default-mode network; SC = subcortical network; SM = sensorimotor network; VIS = visual network.

**Figure S2.** Tested cross-lagged panel models

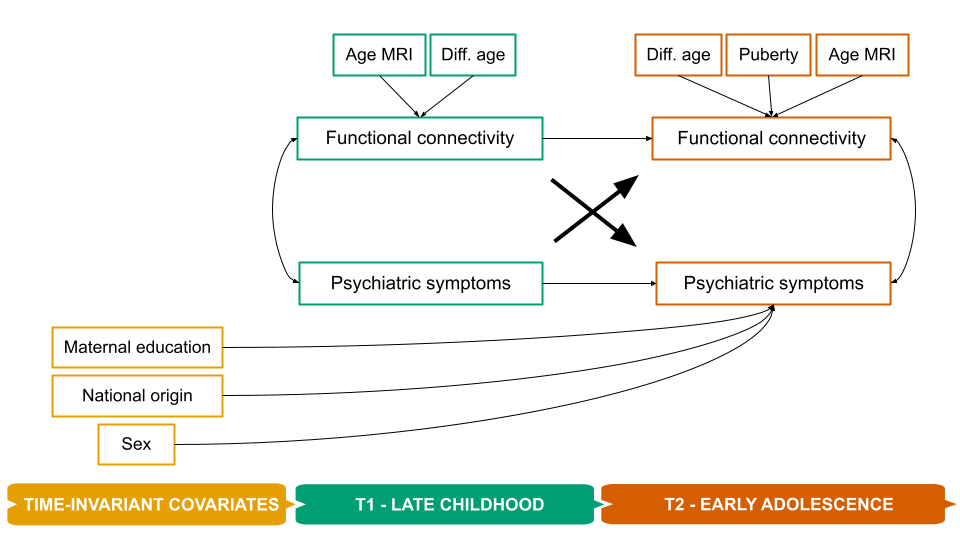

*Note*. Diff = difference; MRI = magnetic resonance imaging. Bolded and larger paths represent the lagged paths of interest. Of note, the difference in age refers to the difference between the age of assessment of the MRI and behavioral measurements.

**Figure S3.** Distribution of static functional network connectivity variables

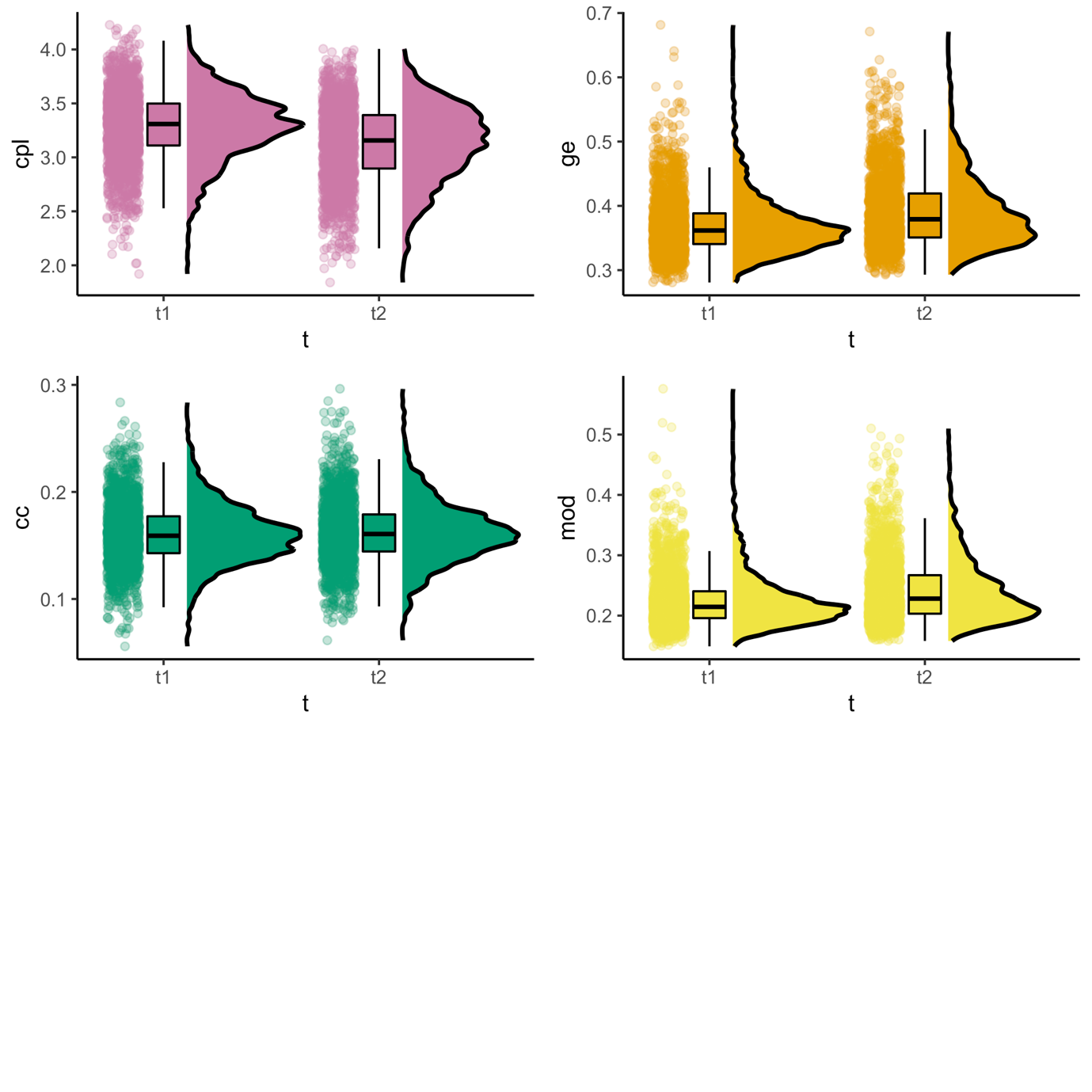

*Note*. Cc = clustering coefficient; cpl = characteristic path length; ge = global efficiency; mod = modularity; t = time-point. Raincloud plots for static functional network connectivity measures across time-point (t1 = age 10 assessment; t2 = age 14 assessment).
